# Modeling polygenic embryo screening in real-world IVF patients demonstrates limitations on efficacy

**DOI:** 10.64898/2026.04.16.26351002

**Authors:** Liraz Klausner, Elvezia Maria Paraboschi, Francesca Mulas, Ludovica Picchetta, Christian Simon Ottolini, Ateret Revital, Danilo Cimadomo, Alberto Vaiarelli, Todd Lencz, Antonio Capalbo, Shai Carmi

**Author notes:** Correspondence (AC), (SC).

## Abstract

**Background:** Polygenic embryo screening (PES) has recently become available to in-vitro fertilization (IVF) patients, allowing them to evaluate the genetic risk of each of their embryos for polygenic conditions such as heart attack or diabetes. Initial modeling predicted that transferring the embryo with the lowest genetic risk for one or more diseases would substantially reduce prevalence in the next generation, with relative risk reductions up to 50%. However, these models assumed the availability of a prespecified number of embryos and that the embryo with the most favorable polygenic risk is born once transferred to the uterus. In reality, a large percentage of embryo transfers do not lead to live births, and IVF frequently results in no or only a single live birth.

**Methods:** To quantify the expected risk reduction in the context of IVF, we used two datasets: 6944 ovarian stimulation cycles from 4452 Italian infertility patients and 2138 stimulation cycles of egg donors. In both datasets, we simulated the hypothetical application of PES in these cycles by assigning patients and their embryos randomly drawn polygenic risk scores for a given disease, assuming that embryos have been transferred in increasing order of their risk, and tracing their birth outcomes. We then compared the risk of the embryo born after hypothetical PES to the risk of an embryo born without PES. We either considered only completed cycles or integrated over possible birth outcomes of non-transferred embryos in incomplete cycles.

**Results:** In stimulation cycles in infertility patients in which all embryos have been transferred and at least one child was born, we estimate that PES will result in relative risk reductions of just ≈1-3%. In an intention-to-screen analysis of all completed cycles (regardless of birth outcomes), relative risk reductions are under 0.5%. The risk reductions increase, as expected, with more euploid blastocysts and with younger maternal age. Including incomplete cycles (in which not all embryos have been transferred) increases risk reductions to ≈2-5%, due to the availability of more euploid blastocysts and a higher live birth rate per transfer in these cycles. Pooling all embryos from all cycles of the same patient increases risk reductions to ≈5-10%. Relative risk reductions in egg donor cycles reach ≈20% even with a single stimulation cycle per donor.

**Conclusions:** With the exception of particularly good-prognosis patients or cycles, typical infertility patients would benefit little from PES. In fertile patients, as represented by egg donors, PES is predicted to achieve greater relative risk reductions. However, even though these reductions are still substantially lower than prior estimates that did not account for realistic live birth rates. Ethical, social, and clinical issues associated with offering PES in the general population should be prioritized in future research.

## Introduction

In preimplantation genetic testing (PGT), clinics or prospective parents evaluate the genetic composition of in-vitro fertilization (IVF) embryos to decide which embryo to use and in what order. The new generation of PGT methods can create genome-wide data for each embryo, allowing clinics to estimate the embryos’ risk of *polygenic* conditions, such as heart attack, breast cancer, type 1 or type 2 diabetes, Crohn’s disease, and schizophrenia (Capalbo et al., 2024; Kumar et al., 2022; Lencz et al., 2021; Li et al., 2024; Siermann, Vermeesch, et al., 2024; Treff, Zimmerman, et al., 2019). This technology, called polygenic embryo screening (PES) or PGT-P, is associated with multiple ethical and social problems, in addition to possible harms to patients (Capalbo et al., 2024; Griffin & Gordon, 2023; Lazaro-Munoz et al., 2021; Polyakov et al., 2022; Siermann, Valcke, et al., 2024; Treff et al., 2022; Turley et al., 2021). Indeed, several professional societies published statements discouraging its clinical implementation (Capalbo et al., 2024; Forzano et al., 2022; Grebe et al., 2024; Klipstein et al., 2026; Lencz et al., 2022).

The main theoretical benefit of PES is reduction in the disease burden in children born after screening. To evaluate the potential risk reduction, studies so far have used statistical modeling, embryo genome simulations, and biobank sibling pair analyses (Capalbo et al., 2024). These studies converged to the conclusion that selecting the embryo with the lowest polygenic risk score (PRS) for a target disease or a disease combination can substantially reduce risk (Cordogan et al., 2025; Klausner et al., 2025; Lencz et al., 2021; Moore et al., 2025; Treff, Eccles, et al., 2019; Treff et al., 2020; Turley et al., 2021; Widen et al., 2022). For example, for diseases such as schizophrenia or Crohn’s disease, the lowest risk embryo out of five is expected to have approximately 50% lower risk compared to a randomly selected embryo (Klausner et al., 2025; Lencz et al., 2021). The large relative risk reductions, despite the generally low accuracy of polygenic risk scores (Sud et al., 2023; Wald & Old, 2019), occur because even a small reduction in genetic liability (risk) can shift an individual away from the high-risk regime where diseases manifest (Lencz et al., 2021).

While promising at face value, these published risk reduction estimates are based on the assumption of the availability of 2-5 viable embryos, all of whom would be born if transferred. However, in realistic IVF cycles (i.e., a single cycle of ovarian stimulation, oocyte retrieval, and fertilization) of human infertility patients, the number of euploid embryos is typically small, particularly with advanced maternal age (Demko et al., 2016; La Marca et al., 2022), and a large proportion of euploid embryo transfers do not result in a live birth (Cimadomo et al., 2023; Tiegs et al., 2021). A recent study estimated the proportion of cycles that would result in at least two live births (the minimum number required for PES) as only 27% (Huttler et al., 2025). While PGT-M patients (testing for monogenic conditions) usually do not suffer from fertility problems, some of their embryos can be carriers of pathogenic variants and thus unavailable for PES, again reducing the pool of available embryos (Lencz et al., 2026). On the other hand, IVF patients using donated eggs, or healthy individuals not suffering from infertility, may have more available embryos. However, even in donor cycles, live birth rate per transfer is only around 50% (Awadalla et al., 2022; Clarke et al., 2025; Insogna et al., 2021; Kushnir et al., 2014; Lindner et al., 2025).

To date, no study has evaluated the utility of PES under realistic IVF conditions using real-world clinical data. Here, we address this gap by quantifying the expected disease risk reduction achievable through PES in two distinct patient populations: infertility patients undergoing IVF with their own eggs and patients using donated eggs. Our data provides empirically grounded estimates of PES utility across the range of patients for whom this technology is being considered.

## Materials and Methods

### Datasets

#### Infertility patients

We used IVF records and outcomes from 6944 oocyte retrieval cycles from 4452 Italian infertile patients whose embryos were tested in Clinica Valle Giulia – Genera center in Rome, (part of IVIRMA Global Research Alliance), between 2015-2022. All MII oocytes were inseminated by ICSI, all embryos were cultured to blastocysts, all blastocysts were biopsied, and only vitrified-warmed single euploid blastocyst transfers were performed. For some analyses, we excluded 1229 cycles where not all euploid embryos have been transferred, leaving 5715 cycles. For each cycle, the dataset included a (randomized) couple ID, consecutive cycle number per couple, year of treatment, maternal age at treatment, maternal and paternal cause of infertility (not considered in this study), and the number of cumulus-oocyte complexes, blastocysts, and euploid blastocysts. For each euploid blastocyst, information was available on whether the embryo was transferred and, if it was, whether the transfer resulted in live birth.

#### Egg donors

We used IVF records from 3408 cycles from 2683 donors whose embryos were used in 16 IVF clinics in Spain between 2020-2024. We excluded donors who did not donate all of their MII oocytes, underwent more than one stimulation cycle, or whose oocytes were used in double embryo transfers, leaving 2138 donation cycles. For each stimulation cycle, the dataset included a (randomized) donor ID and the number of oocytes retrieved. For each recipient, the dataset included recipient ID, the number of blastocysts obtained, and the outcome of each transfer. Not all blastocysts were necessarily transferred. No PGT-A data was available. There were on average 1.25 recipients per donor. For the purposes of this study, we pooled together all transfers from all recipients of each donor.

#### Ethical approval

For both datasets, an ethics committee and a data protection officer approval were obtained for the pseudonymous analysis of data associated with IVF efficacy, efficiency, and safety.

#### Descriptive statistics

The plots shown in Figures 1 and S1-S4 considered each cycle as independent, including cycles from the same couple. For some of the plots, we stratified the cycles based on the standard SART age groups. Binomial confidence intervals were based on Wald’s method.

**Figure 1.**
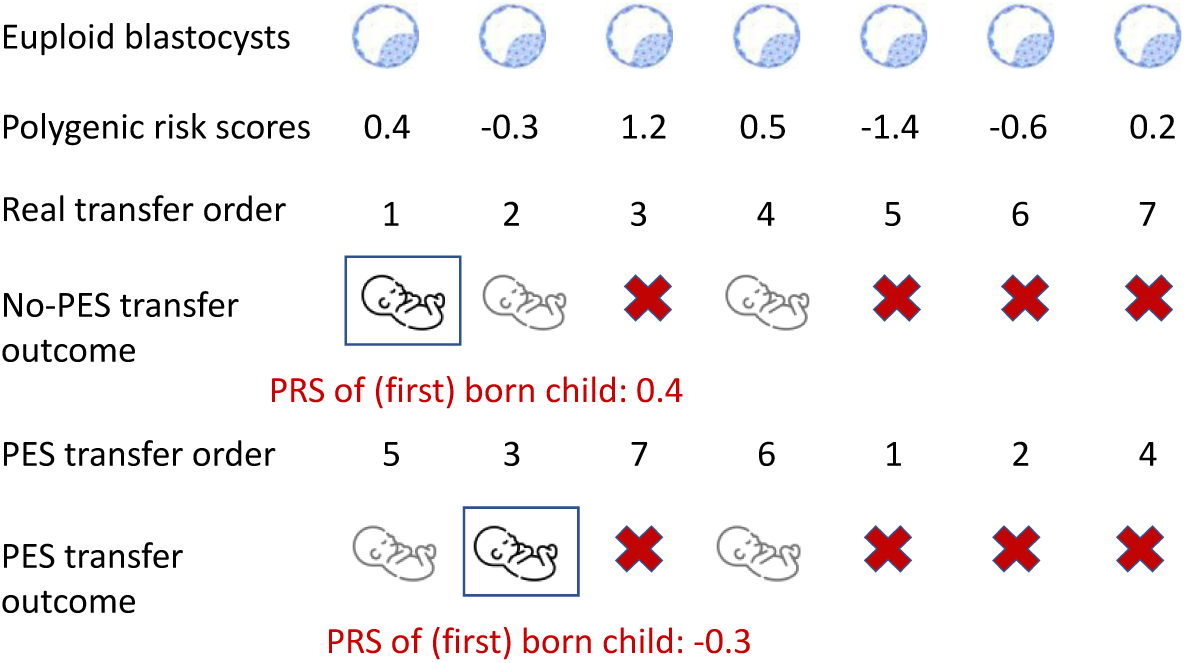
An illustration of embryo prioritization and the risk of the first-born child. We show the PRS for seven hypothetical embryos from the same cycle, all assumed to be euploid. In this example, all seven embryos were transferred and three (the first, second, and fourth) were born. Without PES, the PRS of the first-born child is the PRS of the embryo that was transferred first (0.4 in the example). When simulating PES, we assume that embryo transfer order is determined by the PRS, and that the PRS is uncorrelated with the embryo transfer order. The first two embryos with the lowest PRS (-1.4 and -0.6) were not born. Therefore, the PRS of the first-born child is that of the embryo with the third-lowest PRS (here, -0.3).

### Simulation of polygenic embryo screening

#### Filtering of cycles

For the primary analyses, we considered 930 cycles in which all embryos have been transferred and that resulted in at least one live birth. No additional filtering was performed.

#### Generating PRS for patients and embryos

We assume embryo screening for a single, prespecified disease. Following our published framework based on the liability threshold model (Lencz et al., 2021), we assumed that the disease has an underlying continuous liability with a standard normal distribution. The PRS has a normal distribution in the population with variance *r*^2^, which is also the proportion of variance in liability it explains. Therefore, the PRS of each prospective parent was drawn from a *N*(0, *r*^2^) distribution. We assumed no correlation between the maternal and paternal PRS. Denoting the average parental PRS as *c*, we draw the PRS of each embryo, independently of the other embryos, as *N*(*c*, *r*^2^/2) (Lencz et al., 2021).

#### Estimating the risk of an embryo given its PRS

For disease prevalence *K*, the liability threshold model asserts that individuals are affected if their overall liability exceeds the disease threshold *t* = ϕ^−1^(1 − *K*), where ϕ(⋅) is the cumulative distribution function of the standard normal distribution. The liability of a given embryo can be written as *Y* = *PRS* + *e*, where *e* is the sum of all environmental, random, and genetic risk factors not captured by the PRS. We assumed that *e* and the PRS are uncorrelated, and thus, *e* ∼ *N*(0,1 − *r*^2^). Therefore, the risk of an embryo is 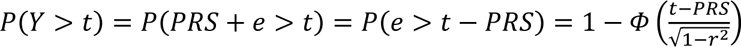 . Note that this holds regardless of the correlation between the values of *e* of different embryos.

#### Simulating embryo prioritization strategies

We computed the risk of the first-born child to be affected either under the baseline model (random selection) or under PES (Figure 1). For random selection, we also assumed that embryo PRS is uncorrelated with morphology or any other factor that would affect transfer order. Therefore, it is sufficient to draw a single PRS to be assigned to the born child. For PES, we again assumed no correlation between PRS and transfer order (but see the sensitivity analyses below). To simulate PES, we drew a PRS for each embryo, sorted the embryos in increasing PRS order, and recorded the PRS of the first embryo that was born based on the real transfer outcomes (Figure 1). In practice, this is equivalent to only drawing PRS for born embryos and recording the lowest PRS (Figure 1). Given the PRS of the born embryo (under either strategy), we computed its risk as described above.

#### Exact risk estimation

The simulation approach is straightforward; however, it results in noisy risk estimates due to the random sampling of parental and embryo PRS. For a fixed number *n* ≥ 1 of live births, Eq. (20) from (Lencz et al., 2021) provides the exact risk of the child with the lowest PRS among all those born,

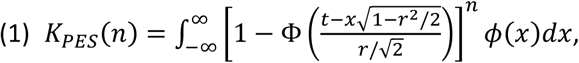

where *t* = Φ^−1^(1 − *K*) is the disease threshold defined above, *r*^2^ is the variance of the PRS, and ϕ(⋅) is the density of a standard normal variable. Eq. (1) integrates over all possible values for the parental and embryo PRS. In our data, the number of live births varies across cycles. We computed the average risk in the patient population as

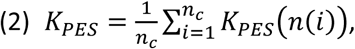

where *n*_*c*_ is the number of cycles with at least one live birth and *n*(*i*) is the number of live births in cycle *i*. Thus, Eq. (2) uses the empirical distribution of the number of live births per cycle and averages the risk of the selected embryo over all cycles. We evaluate the integral in Eq. (1) numerically using R’s built-in *integrate* function.

For random embryo selection, the exact risk, when averaged over all possible PRS, is simply the disease prevalence *K*. We validated that the exact risk estimation (for either selection strategy) gives identical results to the simulation approach.

#### Computing the relative risk reduction (RRR)

Given the average risk of the born embryo with the lowest PRS (*K_PES_*; Eq. (2)) and the risk of a randomly born embryo (*K*), we define the relative risk reduction as

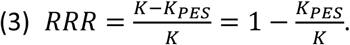

Thus, the RRR is the decrease in average risk (across the entire patient population) between PES-based selection and random selection, scaled by the random selection risk.

We computed 95% confidence intervals for the relative risk reduction (everywhere in the paper) by bootstrapping, resampling cycles with replacement 1000 times and using the empirical [2.5,97.5]% percentiles.

#### An intention-to-screen analysis

The previous analysis might have overestimated the risk reductions, because it only considered cycles with at least one live birth. However, it is also critical to consider an “intention-to-screen” analysis, where the relative risk reduction is defined as zero in case of IVF failure. This definition effectively sets the risk of the future child to the population prevalence *K*. Denote by β the proportion of cycles with no live birth, by *K*_*PES*,≥1_ the risk of a child born after PES for cycles with at least one birth (as determined by Eq. (2)), and by *K*_*PES*,≥0_ the risk after PES for any cycle. We thus have *K*_*PES*,≥0_ = β*K* + (1 − β)*K*_*PES*,≥1_. The relative risk reduction for cycles with at least one live birth is *RRR*_≥1_ = 1 − *K*_*PES*,≥1_/*K*. Thus, *K*_*PES*,≥1_ = *K*(1 − *RRR*_≥1_), or *K*_*PES*,≥0_ = β*K* + (1 − α)*K*(1 − *RRR*_≥1_) = *K*(1 − *RRR*_≥1_) + β*KRRR*_≥1_ = *K*[1 − (1 − β)*RRR*_≥1_]. The relative risk reduction for any cycle is *RRR*_≥0_ = 1 − *K*_*PES*,≥0_/*K* = 1 − [1 − (1 − β)*RRR*_≥1_] = (1 − β)*RRR*_≥1_. Thus, the relative risk reduction in the intention-to-screen analysis is simply the RRR from cycles with at least one live birth, scaled by the proportion of such cycles. For our database, 1 − α = 0.16.

### Additional analyses

#### Transfer order is correlated with PRS

Our primary analyses assumed that the PRS is uncorrelated with the recorded transfer order. However, if embryos with low PRS are likely to be transferred first, and if early transfers have a higher success rate (Figures S3B and S4B), then the risk of the born child will be lower compared to the case of uncorrelated PRS and transfer order (Figure S9). To study risk reductions in this setting, we assumed that the real transfer order is equal to the order of increasing PRS (Figure 1B), and we used the PRS simulation approach. After drawing PRS and sorting the embryos by PRS, we recorded the PRS of the first embryo that was born (based on the actual IVF records) and computed the risk as above. To provide an upper bound on the RRR, we assigned the baseline risk to the disease prevalence *K*, i.e., still assuming no correlation between PRS and transfer order.

#### Including cycles with non-transferred embryos

“Open” cycles with non-transferred embryos in our database have a higher live birth rate per transfer compared to completed cycles (Figures S3 and S4), which may be due to parents discontinuing IVF once reaching their desired family size. Therefore, excluding open cycles may artificially reduce the number of births in the PES simulation (e.g., as observed in (Huttler et al., 2025)) and underestimate the risk reduction. To account for this possible bias, we expanded the primary analysis to all cycles, by integrating over all possible outcomes for the non-transferred embryos. Specifically, we assumed that each non-transferred embryo is born with probability *p*_*birt*ℎ_, which we estimated based on data from *transferred* embryos in either completed cycles only (*p*_*birt*ℎ_ = 0.39), the entire data (*p*_*birt*ℎ_ = 0.48), or open cycles only (*p*_*birt*ℎ_ = 0.66). We then extended Eq. (2) as follows,

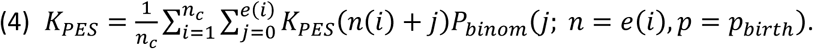

In Eq. (4), for cycle *i*, *n*(*i*) is the number of births from *transferred* embryos, *e*(*i*) is the number of *non-transferred* embryos, 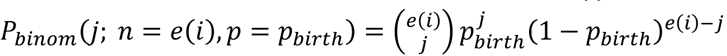 is the binomial probability for *j* live births to result from hypothetical transfers of the remaining *e*(*i*) embryos, and *K*_*PES*_(*n*) is computed as in Eq. (1). In other words, for each open cycle, we sum over all possible numbers of births that may result from the non-transferred embryos, and then compute the risk given selection of the child with the lowest PRS from among all embryos born in that cycle. For completed cycles, *e*(*i*) = 0, and the equation reduces to Eq. (2). We then compute the RRR as in Eq. (3).

In Eq. (4), we account for cases of no birth (terms in the sum with *n*(*i*) = *j* = 0) in two ways. To perform an “intention to screen” analysis, the external sum is over all *n*_*c*_ = 6944 cycles in the database, and for terms in which *n*(*i*) = *j* = 0, we set *K*_*PES*_(0) = *K*. To estimate the risk reduction when conditioning on at least on birth, we modify Eq. (4) as follows. First, we continue to sum over all cycles, but we set *K*_*PES*_(0) = 0. Second, we scale *K*_*PES*_ by the expected proportion of cycles with at least one birth. This results in the following equation,

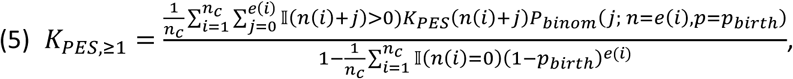

where ‖(*x*) is the indicator function (‖(*x*) = 1 if *x* is true and ‖(*x*) = 0 otherwise).

#### Aggregating over all cycles from the same couple

We simulated the case when patients pool embryos from multiple cycles before ranking them for transfer based on PRS. We used the entire dataset (6944 cycles), integrating over all possible birth outcomes as in the case with leftover embryos. For each couple (same couple ID), we used all cycles regardless of calendar year or outcome. We assumed no correlation between PRS and transfer order. We then used Eqs. (4) and (5) to compute the risk reduction for the intention-to-screen analysis and when conditioning on at least one birth, respectively. Note that in Italy, new cycles can be initiated only once all embryos from an existing cycle have been transferred.

#### Egg donor cycles

To predict the outcome of non-transferred embryos, we estimated the live birth rate using all transfers in the egg donor datasets, resulting in *p*_*birt*ℎ_ = 0.33. This rate is lower than in the infertility database, because the embryos derived from the donated eggs were not tested with PGT-A and thus some of them have likely been aneuploid. The live birth rate was approximately constant as a function of age (Figure S6D; range 18-35). We then used Eq. (4) to predict the risk with PES in an intention-to-screen analysis and Eq. (5) when conditioning on having at least one live birth. For each donor, each cycle in our analysis corresponds to a single stimulation cycle, and we pooled outcomes from all generated blastocysts regardless of the ID of the recipient.

## Results

### The IVF dataset

We used records from 6944 cycles of 4452 IVF patients, all with an infertility/subfertility indication. The mean and SD of the (female) patient age was 39 ± 3.42 years. In Figure 2, we plot the average number of oocytes, blastocysts, euploid blastocysts, and live births per cycle vs the maternal age. Notably, the mean number of euploid blastocysts per cycle was only 0.88 and the mean number of live births was 0.17. In Figures S1-S4, we show, respectively, the distributions of the variables reported in Figure 2 across all cycles; the distribution of the number of cycles and the maternal age per couple; and the average live birth rate per euploid embryo transfer vs the maternal age, the transfer number within the cycle, and the year of treatment (across all or only completed cycles). Overall, the analysis demonstrates the expected pattern of a “leaky pipeline”, whereby the number of live births per cycle can be very small despite larger numbers of oocytes or blastocysts, as well as the decline in embryo and live birth count with maternal age.

**Figure 2.**
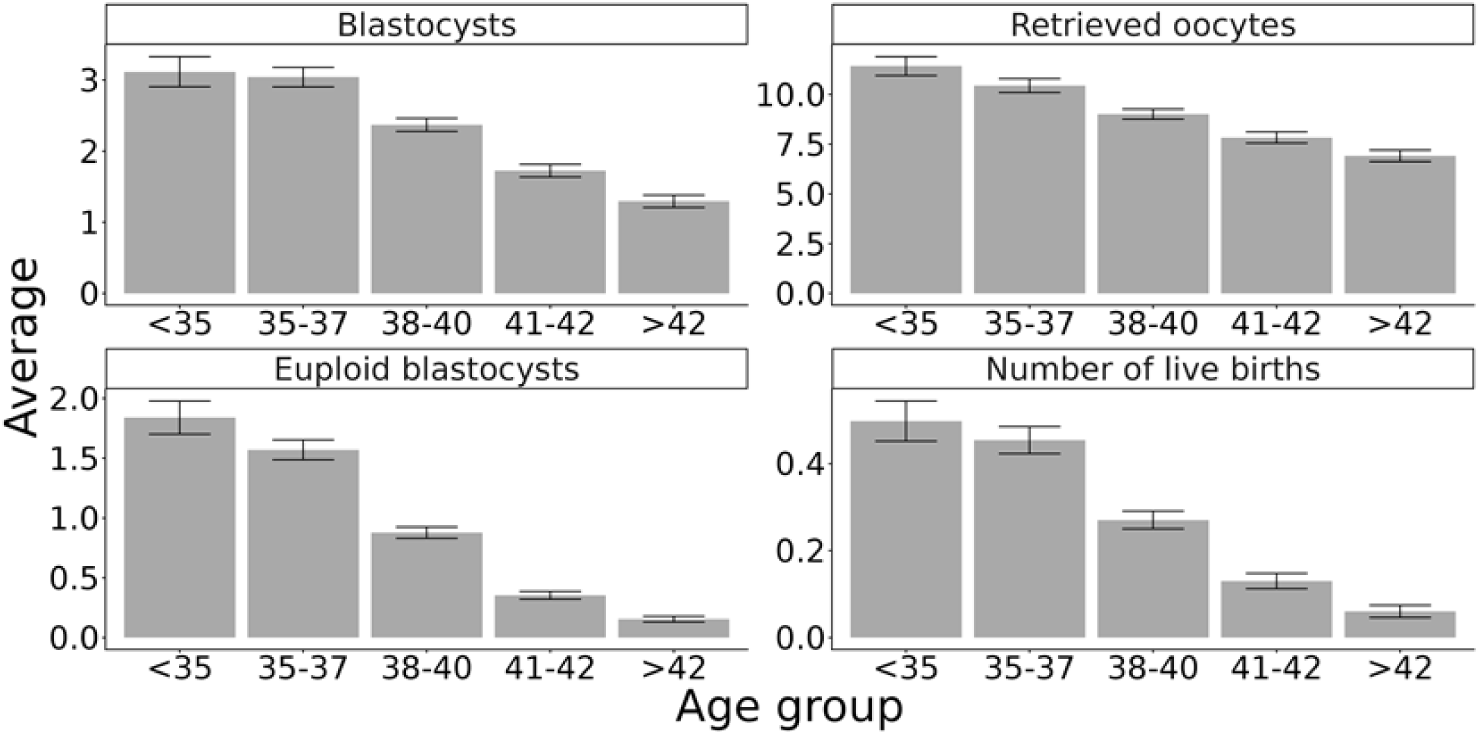
The average number of oocytes, blastocysts, euploid blastocysts, and live births per cycle vs the maternal age. The averages are over all 6944 cycles, except the number of live births, where it is over the 5715 cycles for which all embryos have been transferred. Bars denote 95% confidence intervals for the mean.

### Simulating polygenic embryo screening

We assumed screening for a single, prespecified disease. To predict risk reduction due to PES, we assigned randomly-generated PRS values to patients and simulated the PRS transmitted to each embryo. We then computed the risk of each embryo based on the liability threshold model (Methods). We initially only considered the 5715 cycles in which all embryos have been transferred.

To simulate PES, we assumed that embryos were transferred in an increasing order of their PRS, and we recorded the PRS and the disease risk of the first transfer that resulted in a live birth, based on the real documented IVF outcomes (Methods; Figure 1). To reduce the simulation noise, we used equations that analytically integrate over all possible PRS (Methods). For comparison, the risk of the first-born embryo when the transfer order is random with respect to the PRS is the disease prevalence (Methods). We computed the relative risk reduction (RRR) based on the difference between the PES-based risk and the random transfer risk, each averaged over all cycles (Methods).

In Figure 3A, we plot the RRR for a number of realistic values for the disease prevalence *K* and the PRS accuracy *r*^2^(the proportion of disease liability explained by the PRS; (Capalbo et al., 2024)). We initially focused only on the 930 cycles with at least one live birth, reasoning that RRR is not well defined when no child is born. Across all parameters, RRR is only ≈1-3%, substantially lower than the previously predicted risk reductions that assumed 2-5 births per cycle. For example, for type 2 diabetes, with prevalence around 10%, the predicted RRR with PES was only 1.4%, compared to previously estimated 22% with two born embryos and 41% with five (Capalbo et al., 2024; Lencz et al., 2021). For a lower prevalence disease such as schizophrenia (prevalence around 1%), the RRR was 2.1%. Finally, for an even rarer disease such as multiple sclerosis (prevalence around 0.5% (Nielsen et al., 2005; Shams et al., 2023)), the RRR was 2.2% (assuming *r*^2^ = 0.1 for all conditions).

**Figure 3.**
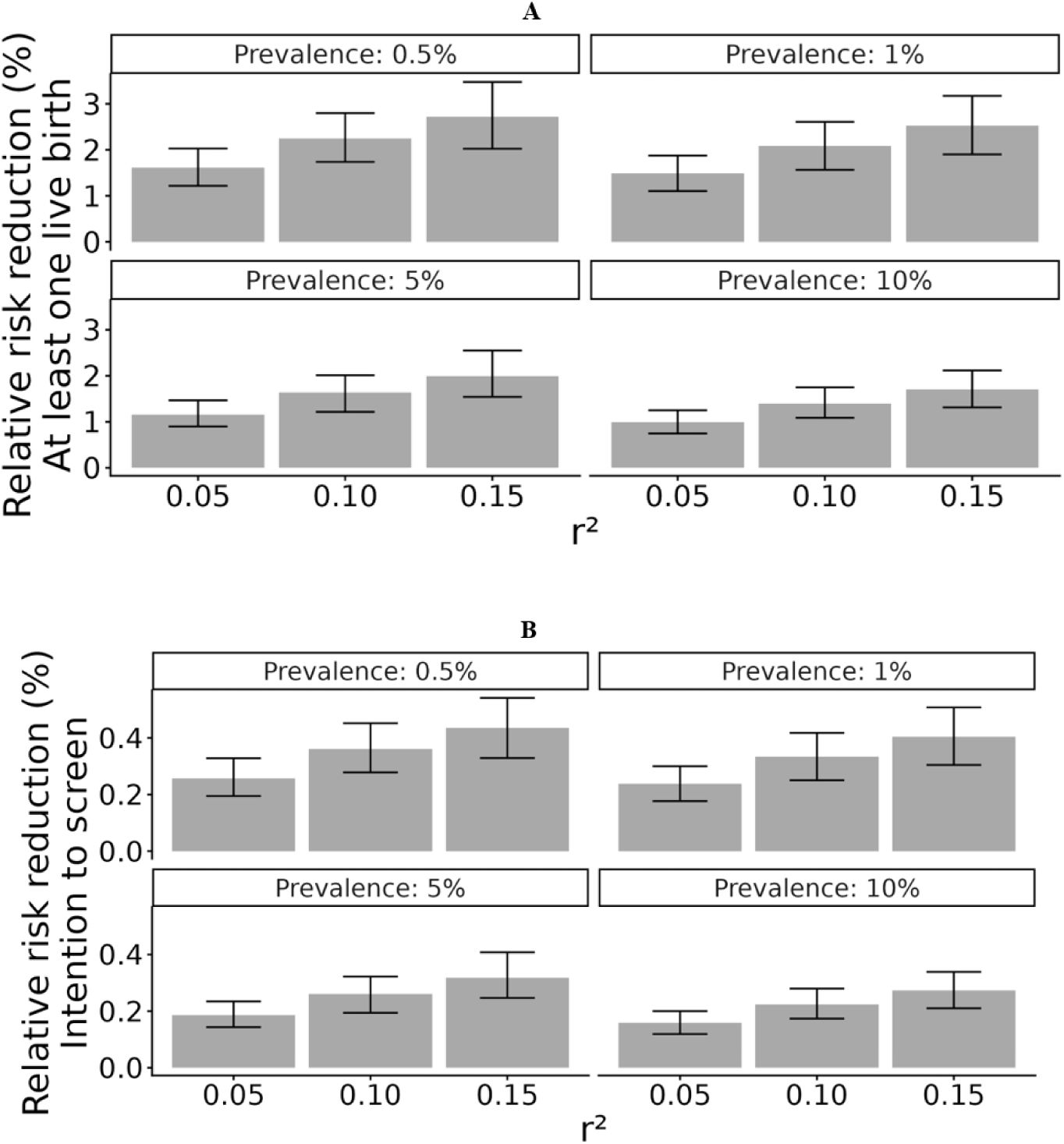
The predicted risk reduction when selecting an embryo for transfer based on having the lowest polygenic risk score for a target disease. We plot the relative risk reduction (RRR) vs the PRS accuracy *r*^2^(the proportion in disease liability explained by the PRS) for a few values of the disease prevalence (see legend). The estimates in panel (A) are based on 930 cycles in which all embryos have been transferred and that have resulted in at least one live birth. The risk of the selected embryo was computed based on the liability threshold model (Methods). We computed the RRR by comparing the risk of the (born) embryo with the lowest (simulated) PRS to the risk of a randomly selected (born) embryo. In panel (B), we show the “intention-to-screen” estimates based on 5715 cycles in which all embryos have been transferred. Here, we set the RRR to zero in case the cycle has resulted in no live births. The bars here and in all other figures showing risk reductions denote 95% confidence intervals, obtained by bootstrapping over cycles.

It can be argued that in cycles in which no child was born, the risk of any future child is no different than the population risk, and thus the RRR should be defined as zero. In this “intention-to-screen” type of analysis, which is shown in Figure 3B, risk reductions are even lower and are now under 0.5% across all parameters.

PES-based risk reduction strongly depends on the number of euploid embryos, and, consequently, on the maternal age. For fixed typical values of PRS accuracy and disease prevalence (*r*^2^ = 0.1, *K* = 1%), we plot in Figures 4A and 4B the risk reduction vs the maternal age and the number of euploid blastocysts, respectively. The risk reduction increased from 7.3% with 2 available euploid blastocysts to 9.4% with 4. Accordingly, risk reduction is greater for younger patients, ranging from 2.8% for females under 35 to 0.5% for patients over 42.

**Figure 4.**
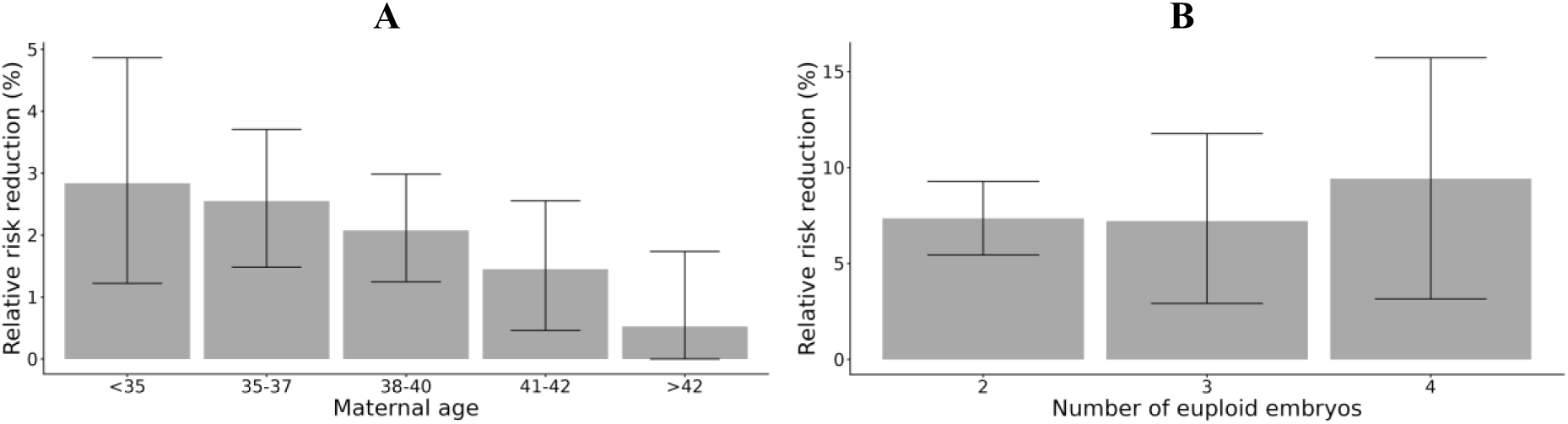
The predicted risk reduction vs the maternal age (A) and the number of euploid embryos (B). We used the same procedure as in Figure 3A, namely the plots are based on 930 cycles in which all embryos have been transferred and at least one was born. The PRS accuracy was set to *r*^2^ = 0.1 and the disease prevalence to *K* = 1%. Note that with a single euploid embryo, the risk reduction is by definition zero. Even with two or more euploid embryos, given that the selected embryo may not be born, risk reduction is not guaranteed.

### Sensitivity analyses

To assess the robustness of our results, we considered the following additional analyses.

#### Transfer order

In the above analyses, we generated a random PRS for each embryo, and we examined the transfer outcomes of embryos in increasing order of their PRS (Figure 1). This implicitly assumed no correlation between PRS and embryo competence, in the absence of any data to the contrary. However, if embryos with low PRS tend to be transferred first (e.g., due to having better morphology), and further, if earlier transfers have better outcomes (Figures S3B and S4B), this would lead to the birth of children with lower risk. This argument is visualized in Figure S9. To predict risk reductions in such a setting, we now assumed that in each cycle, embryo PRS are sorted based on their real transfer order. For conservativeness, we assumed that without PES, there is no correlation between transfer order and embryo PRS. However, as we show in Figure S5, even so, risk reductions are still in the range ≈1-2.5%, with minimal difference from Figure 4A. This may be explained by the large fraction of the cycles with only one euploid blastocyst (Figure S1C), in which transfer order is irrelevant.

#### Including incomplete cycles

Our primary analysis included only “completed” cycles in which all euploid embryos have been transferred, and thus, the transfer outcomes were available for all embryos. However, birth rates in “open” cycles are high, at 66% per transfer (compared to 39% in completed cycles; Figures S3 and S4), possibly because couples who have already given birth after using their high-quality embryos do not transfer their remaining embryos. In addition, open cycles have more euploid blastocysts (mean 2.93) compared to completed cycles (mean 0.44). To predict risk reductions when including all cycles, we integrated over all possible hypothetical transfer outcomes of the non-transferred embryos using birth rates per transfer estimated using either the entire dataset, completed cycles only, or open cycles only (Methods). The results (Figure 5A) show that the RRR (in an intention-to-screen analysis) indeed increases substantially compared to the primary analysis, particularly when estimating birth rates only from open cycles. However, even in this setting, the RRR is only in the range ≈2-6%. When conditioning on at least one birth, the RRR is in the range 10-20% (Figure S6A).

**Figure 5.**
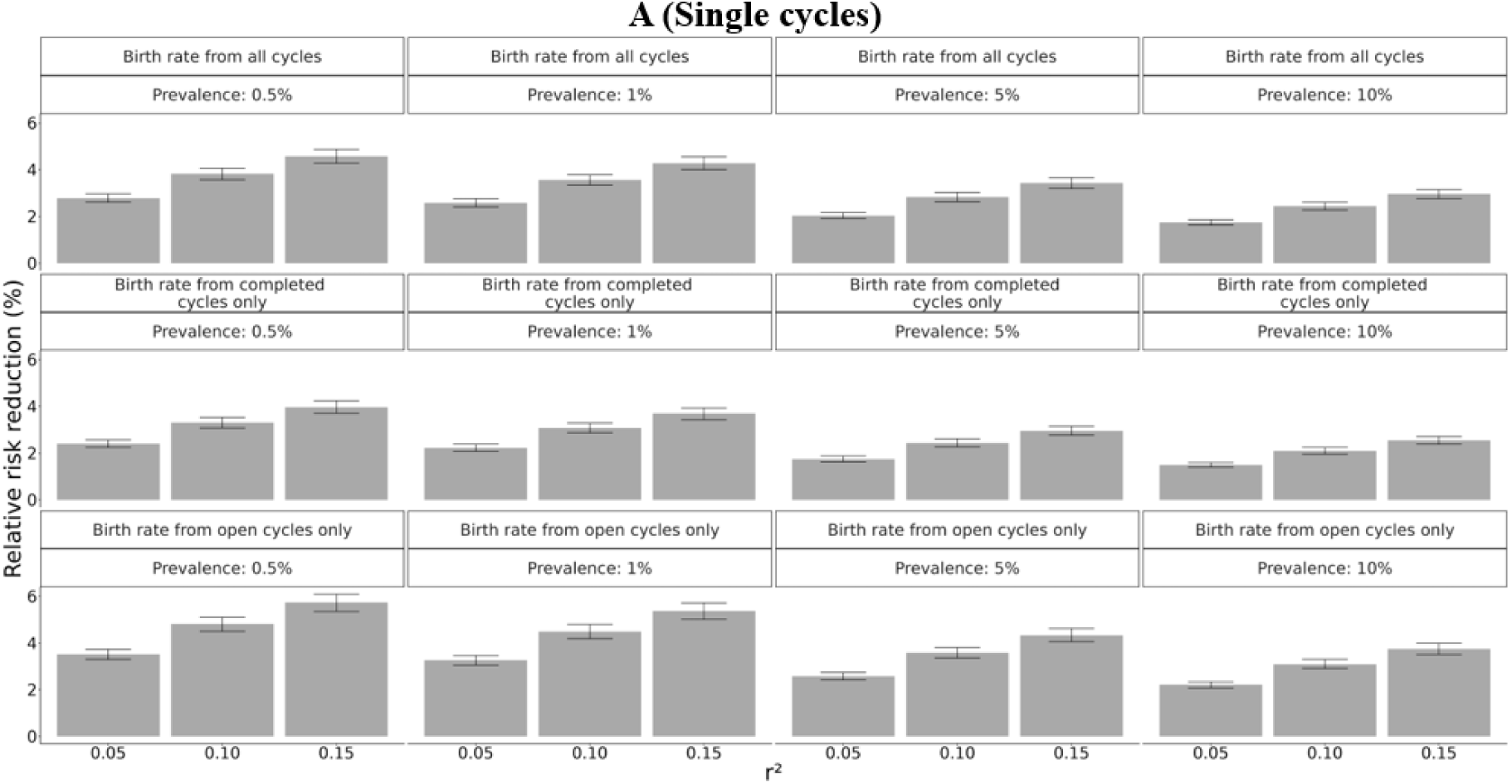
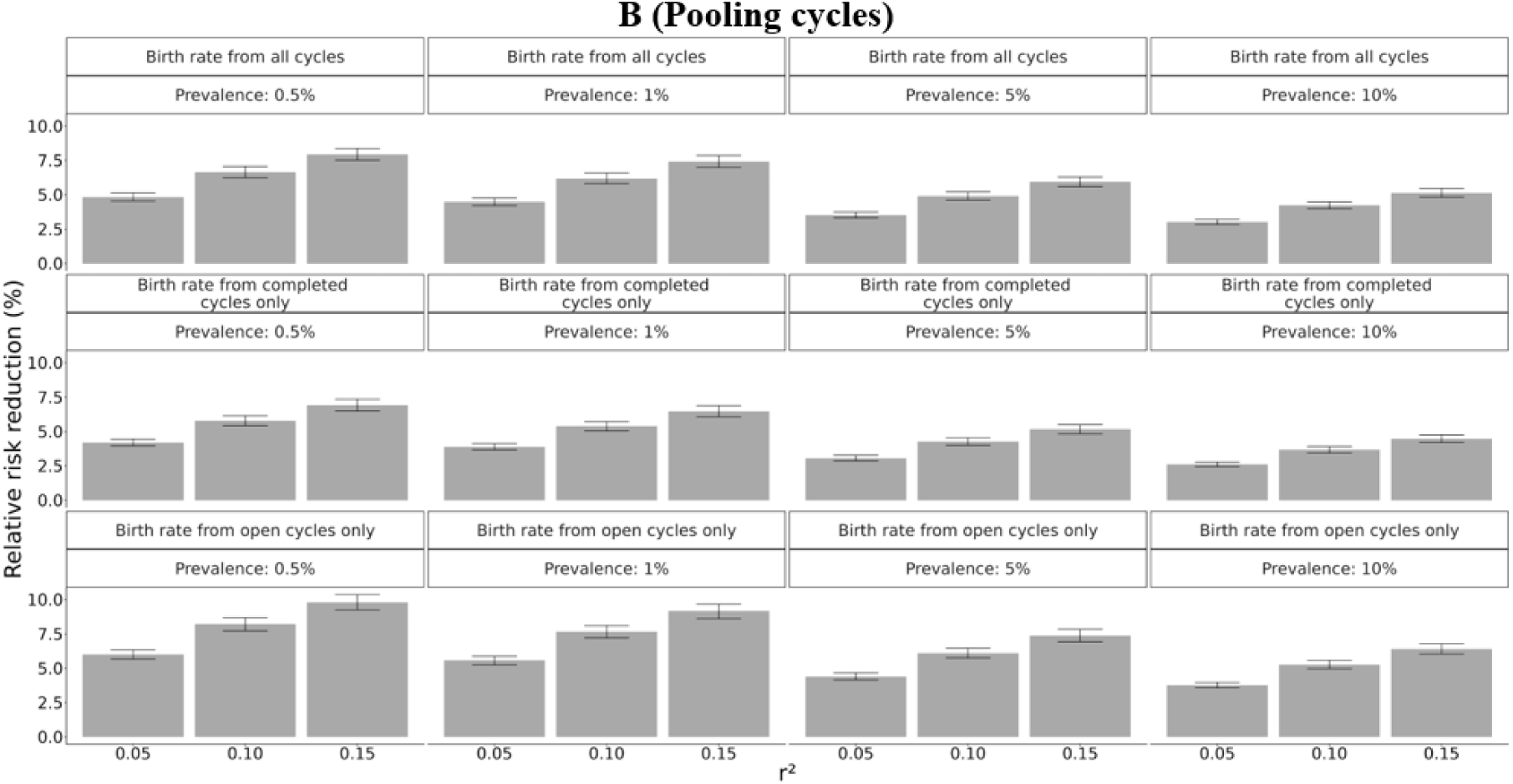
Sensitivity analyses. (A) Risk reductions when including open cycles, i.e., cycles in which not all embryos have been transferred. This analysis is therefore based on all 6944 cycles in the database (assigning zero risk reduction to cycles with no live birth, in an intention-to-screen analysis). We integrated over all possible transfer outcomes (live birth/no live birth) of embryos not transferred, based on a live birth probability estimated using either the entire dataset, completed cycles only (in which all embryos have been transferred), or open cycles only (see legend). (B) Risk reductions when pooling embryos from all cycles of the same couple. Here, we again used the entire dataset (6944 cycles), integrating over the possible birth outcomes for non-transferred embryos as in panel (A).

#### Using all cycles of each couple

Our primary analysis considered a single IVF cycle as the atomic unit for which risk reduction is evaluated. However, it can be argued that patients may generate embryos over multiple IVF cycles before prioritizing them for transfer. To evaluate risk reductions in this setting, we pooled all embryos from all IVF cycles of the same couple, and simulated PES as above. The mean and SD of the number of cycles per couple was 1.56 ± 0.9 (Figure S2). The results (Figure 5B) show, as expected, an increase in the RRR up to ≈5-10% in the intention-to-screen analysis or ≈10-20% when conditioning on at least one live birth (Figure S6B).

### Egg donor cycles

The small risk reductions we showed in our primary analysis are driven by the very small number of live births per cycle that characterize IVF in infertility patients (Figure 2). However, IVF cycles with egg donation have a much better prognosis compared to patients using autologous eggs, given that egg donors are typically younger and have more euploid embryos (Goldman et al., 2017; Rodríguez-Varela et al., 2024). This results in more live births (Kushnir et al., 2014) and is thus expected to result in greater risk reductions.

To predict risk reduction due to PES in egg donation cycles, we used a dataset of 2138 cycles, after excluding donors who did not donate all of their oocytes, underwent more than one stimulation cycle, or whose oocytes were used in double embryo transfers. We pooled all blastocysts from the same donor, regardless of recipient. Figure S7 shows the distribution of the number of oocytes retrieved per cycle, the number of blastocysts per cycle, the donor age, and the live birth rate per transfer vs the donor age. As expected, the average number of blastocysts in these cycles, 5.29, is much larger than for infertility patients (2.26; Figure S1). PGT-A was not performed for blastocysts in this dataset.

We simulated PES as for the infertility patients. Specifically, we included cycles in which not all embryos were transferred and integrated over the possible outcomes of the non-transferred embryos. The live birth rate per blastocyst transfer was estimated as 0.33 based on all transfers. The risk reductions predicted for the egg donor cycles (Figure 6), in an intention-to-screen analysis, are much higher than those of the infertility patients, reaching ≈20% (compare to Figure 3), or even ≈30% when conditioning on having at least one live birth (Figure S8).

**Figure 6.**
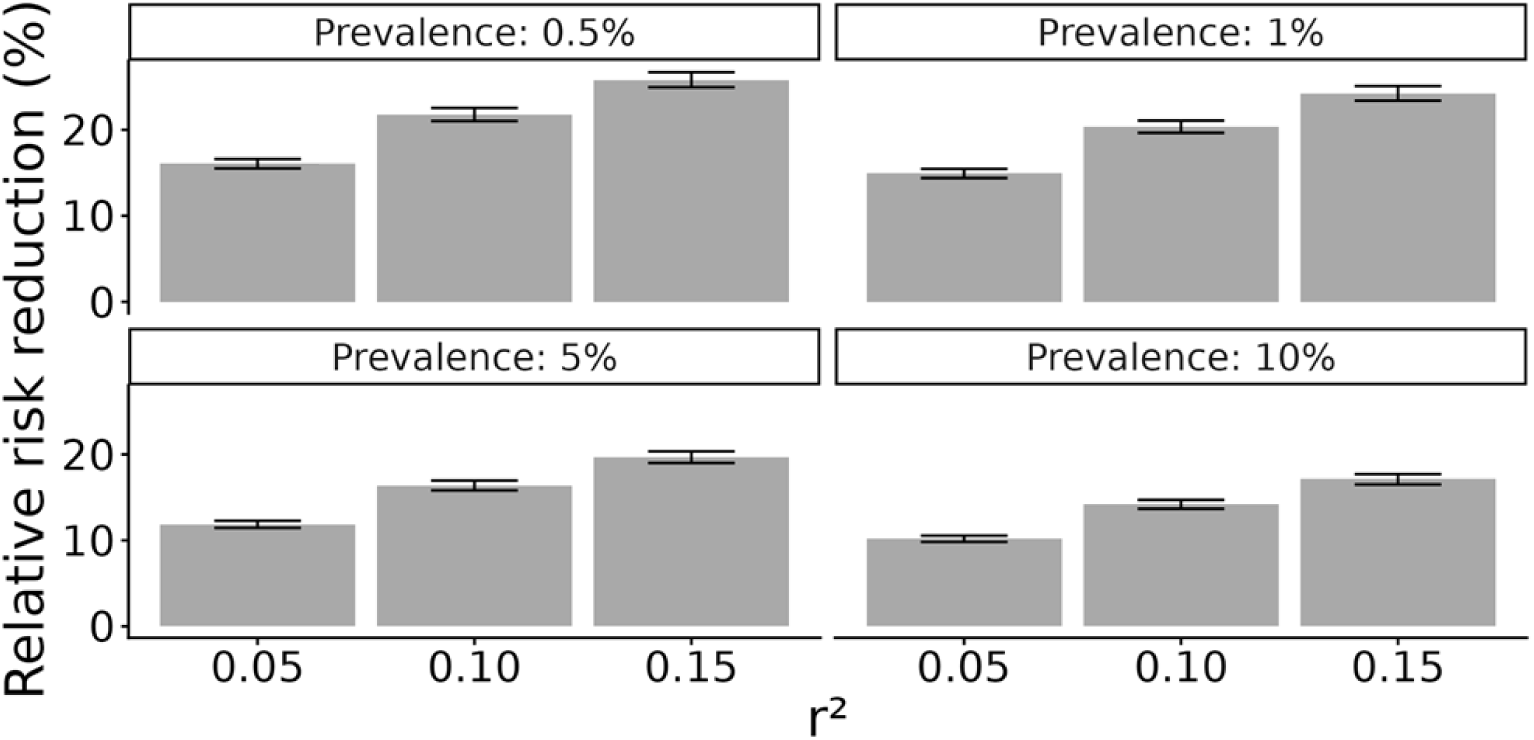
Risk reduction for egg donor cycles. The figure shows the relative risk reduction vs the PRS accuracy *r*^2^for several values of the disease prevalence (see legend). The estimates are based on 2138 cycles and were performed as in Figure 5A, by integrating over the outcomes of non-transferred embryos using the overall live birth rate per blastocyst transfer in these cycles. The results represent an intention-to-screen analysis, where the risk reduction is set to zero in cycles with no live births.

## Discussion

PES was previously predicted to result in major reductions in future disease burden, with some estimates reaching 50% or larger relative risk reductions (Cordogan et al., 2025; Lencz et al., 2021; Moore et al., 2025; Treff, Eccles, et al., 2019; Treff et al., 2020). Here, we predict the utility of PES for real IVF patients, for whom the number of viable embryos and live births may be small. Predicted risk reductions in infertility patients are low: less than 0.5% in an “intention-to-screen” analysis in “completed” cycles (in which all embryos were transferred) and ≈2-5% when including imputed outcomes for non-transferred embryos. Thus, PES has little utility for most infertility patients.

We also identified two settings in which predicted risk reductions are somewhat greater. First, pooling all embryos across all cycles for a given couple increases risk reductions to ≈5-10%. However, patients would have to undergo multiple oocyte retrieval cycles, substantially increasing the cost, inconvenience, and time required to achieve a pregnancy. Further, pooling embryos across cycles is not permitted in some jurisdictions (Vergallo et al., 2025). Second, in egg donor cycles, in which the number of blastocysts is large compared to infertility patients, risk reductions reached ≈20%. However, even this is still substantially less than previous estimates in idealized but unrealistic scenarios, such as having five embryos with birth potential (Cordogan et al., 2025; Lencz et al., 2021; Moore et al., 2025; Treff, Eccles, et al., 2019; Treff et al., 2020). In addition, other limitations remain, such as small absolute risk reductions for the less common conditions (and thus a very large number of patients that need to be screened in order to avoid a single disease case), lower gains in patients of non-European ancestry, and uncertainty regarding PRS accuracy in the future (Capalbo et al., 2024)

The main limitation of our analysis is that our estimates require mathematical models of embryo risk and are thus sensitive to deviations from the models’ underlying assumptions. Specifically, we assumed a normal distribution of the PRS in parents, a normal distribution with half the variance of the PRS in embryos given parental PRS, and we used the liability threshold model to compute the disease risk of an embryo given its PRS. We previously thoroughly analyzed these assumptions and found them to reasonably approximate real data (Lencz et al., 2021). While future studies should explore more nuanced models, it must be emphasized that no real IVF data could directly estimate risk reductions, given that polygenic diseases typically manifest in adulthood or adolescence.

Another key limitation is the large difference between completed cycles (in which all embryos have been transferred) and incomplete (“open”) cycles. The live birth rate per euploid embryo transfer was much higher in open cycles (0.66 vs 0.39; Figures S3B and S4B), raising the question of which live birth rate should be assumed for non-transferred embryos. The reasons for not transferring all embryos likely vary across patients, and while it is plausible that the leftover embryos have lower quality than those transferred, it is difficult to validate such a hypothesis. The number of euploid blastocysts was also much higher in the open cycles (2.93 vs 0.44). In future work, data from healthcare settings in which only a single oocyte is fertilized per stimulation cycle may provide more robust live birth rate estimates.

Finally, another limitation is that our results are based on specific IVF databases. As such, our risk reduction estimates may not generalize to patients in other countries or clinics with a different distribution of indications. Unfortunately, our database was too small for subgroup analyses based on additional patient characteristics. However, given that it is well known that the number of euploid embryos and live births per cycle is small for infertility patients (Goldman et al., 2017; Rodríguez-Varela et al., 2024), we anticipate our qualitative conclusions to remain valid. Finally, our model lacks nuance regarding the willingness of patients to discard embryos and go through additional stimulation cycles for the purpose of PES. Future research on parental decision-making would enable more realistic risk reduction estimates.

Evaluations of PES so far have broadly considered its utility and harms for all patients requiring or who may elect IVF. However, different prospective PES patient groups differ materially in their benefit and risk profiles. In our review paper (Capalbo et al., 2024), we proposed that future studies should consider three patient groups: (1) people already requiring IVF; (2) (fertile) people with a personal or familial polygenic disease history attempting to avoid “transmission” of a *specific* disease to the next generation; and (3) healthy people seeking PES to enhance the health of their children. This work is (to the best of our knowledge) the first attempt to evaluate the utility of PES specifically in two of these patient groups. Our results show divergence between predicted outcomes: in most infertility patients (group (1)) PES provides little benefits; in contrast, our results for the egg donors suggest that PES can lead to non-trivial risk reductions in healthy individuals (group (3)), even if still much smaller than previously predicted.

Despite the minimal clinical utility for infertility patients as a whole, there are a few points to consider. (1) PES could be beneficial for certain patients who have a large number of euploid blastocysts, particularly if only a single birth is desired. (2) PES could be important for patients undergoing PGT-M for moderate penetrance pathogenic variants, as including a PRS in the risk assessment may impact the identity of the lowest-risk embryo (Lencz et al., 2026). (3) PES in infertility patients is associated with less harms compared to PES in patients who do not otherwise require IVF. The main possible harms for infertility patients include (i) an unnecessary embryo biopsy and costly genetic testing; (ii) discarding viable embryos, thereby adversely impacting reproductive goals; (iii) insufficient counseling; and (vi) choice overload (Capalbo et al., 2024). However, these harms are partly avoidable if (i) patients are already undergoing PGT-A, such that embryos are biopsied and sequenced regardless of PES; (ii) patients do not plan to discard any viable embryos (even if that would lead to lower risk reductions); (iii) adequate counseling is provided; and (vi) embryo ranking is performed by labs rather than by patients.

Our results for egg donors suggest that healthy, fertile individuals who can generate a large number of euploid blastocysts could achieve non-negligible risk reductions using PES. While the predicted risk reductions are still smaller than previously reported, and further, risk reductions are limited for a specific disease, the possibility of fertile individuals undergoing IVF only for the purpose of PES raises ethical, social, and clinical concerns (Capalbo et al., 2024). These include commercialization and medicalization of reproduction, market pressures on parents, and the role of healthcare professionals (Pereira et al., 2022). Further, offering PES to the general public raises ethical concerns regarding a slippery slope towards eugenics and selection for physical and cognitive traits (Karavani et al., 2019; Lazaro-Munoz et al., 2021; Siermann, Valcke, et al., 2024; Treff et al., 2022; Turley et al., 2021). While embryo screening for traits is supported by only a minority of the general public (Barlevy et al., 2025; Furrer et al., 2024), it may nevertheless be one day adopted, particularly that it is now offered by two companies (Dwoskin & Torbati, 2025). Additional concerns include the right of the child to an open future, distraction from social and modifiable causes of disease; unequal access and benefits across populations; and future stigmatization against and discrimination of people affected with the conditions screened (Capalbo et al., 2024). Defining the circumstances under which PES could be ethically offered to non-fertility patients (e.g., those with a personal or a family history of a severe polygenic disease) and how it should be implemented should be the focus of future studies.

Finally, given the higher risk reductions predicted here in the egg donor data, an additional concern is that our results may push healthy individuals or IVF patients interested in PES towards using donated eggs. Increasing demand for donated eggs raises multiple ethical concerns (Deng et al., 2025; Marshall, 2017), particularly exploitation of potential donors and commodification of egg donation.

## Acknowledgements

LK, TL, and SC were supported in part by the National Human Genome Research Institute of the National Institutes of Health under award number R01HG011711. The content is solely the responsibility of the authors and does not necessarily represent the official views of the National Institutes of Health. Some of the work on this paper was performed when authors AC, FM, EMP, LP, and CSO were employees of Juno Genetics.

## Code availability

R code that was used to generate risk reduction estimates can be found at https://github.com/Lirazk/IVF.

## Data availability

Due to privacy reasons, patient data cannot be shared.

## Author contribution

Conceptualization: SC, AC, TL; Data curation: EMP, FM, LP; Formal analysis: LK, EMP, FM, LP; Funding acquisition: SC, TL; Investigation: LK, AR, SC; Methodology: LK, SC; Project administration: SC, AC; Resources: DC, AV; Supervision: SC, AC, TL; Visualization: LK, SC; Writing – original draft preparation: SC; Writing – reviewing & editing: SC, LK, AC, DC, CSO, TL.

## Conflicts of interests

EMP, FM, CSO, and AC are employees of Genethyx. SC is a paid consultant at MyHeritage.

---

**Figure S1.**
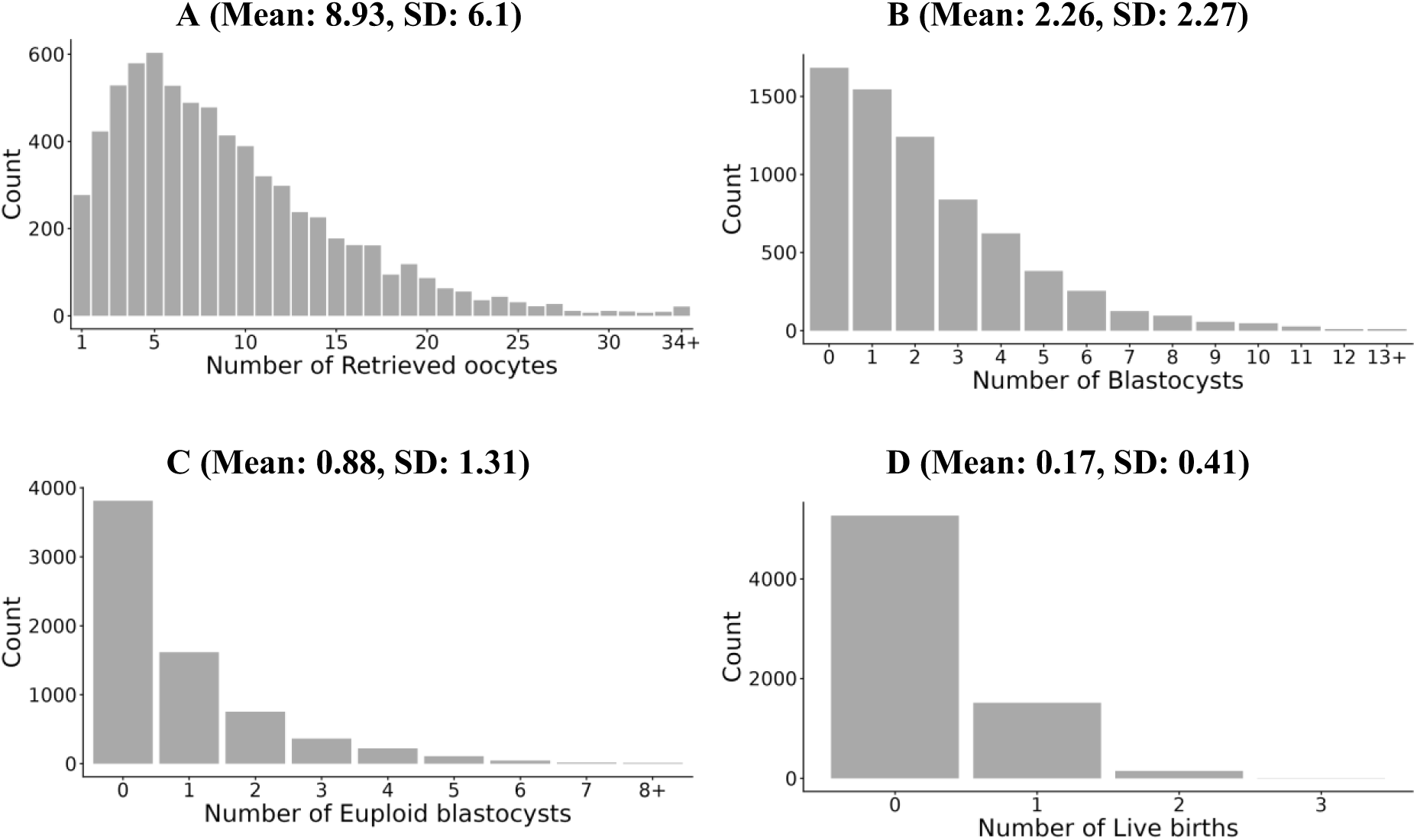
The distribution of IVF outcomes. The figure shows the distribution of the number of oocytes (A), blastocysts (B), euploid blastocysts (C), and live births (D) per cycle. The data includes all 6944 cycles, except the number of births, where it is over the 5715 cycles in which all embryos have been transferred. All panels include all maternal ages.

**Figure S2.**
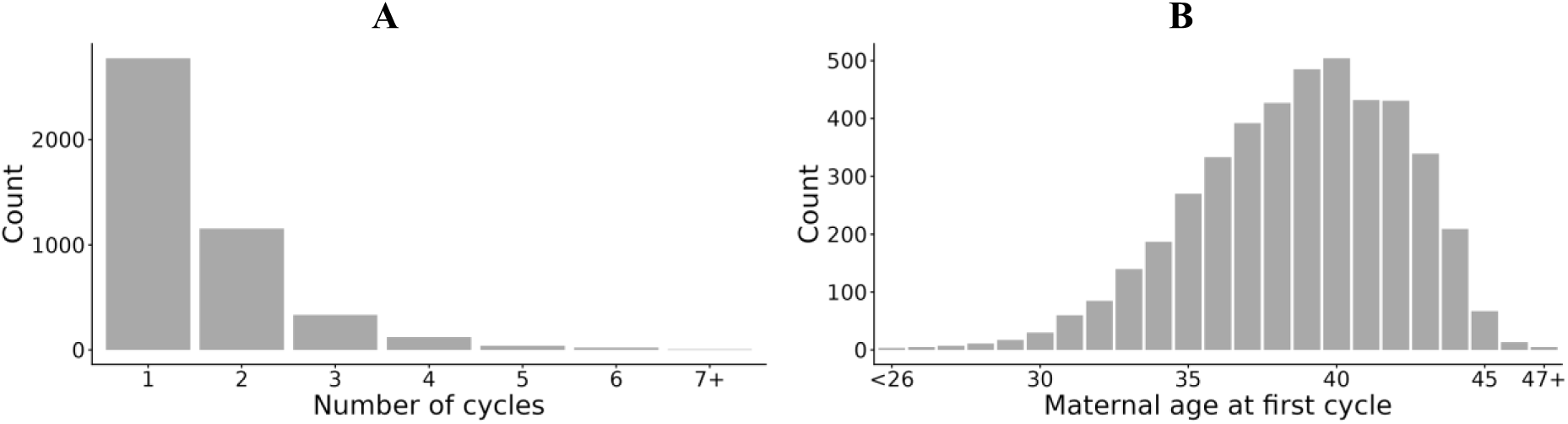
Characteristics of the couples. The figure shows the distribution of the number of cycles (A) and the maternal age (B) per couple. The data includes all 4452 couples regardless of cycle(s) outcome. For couples with multiple cycles, we used the maternal age at the first cycle.

**Figure S3.**
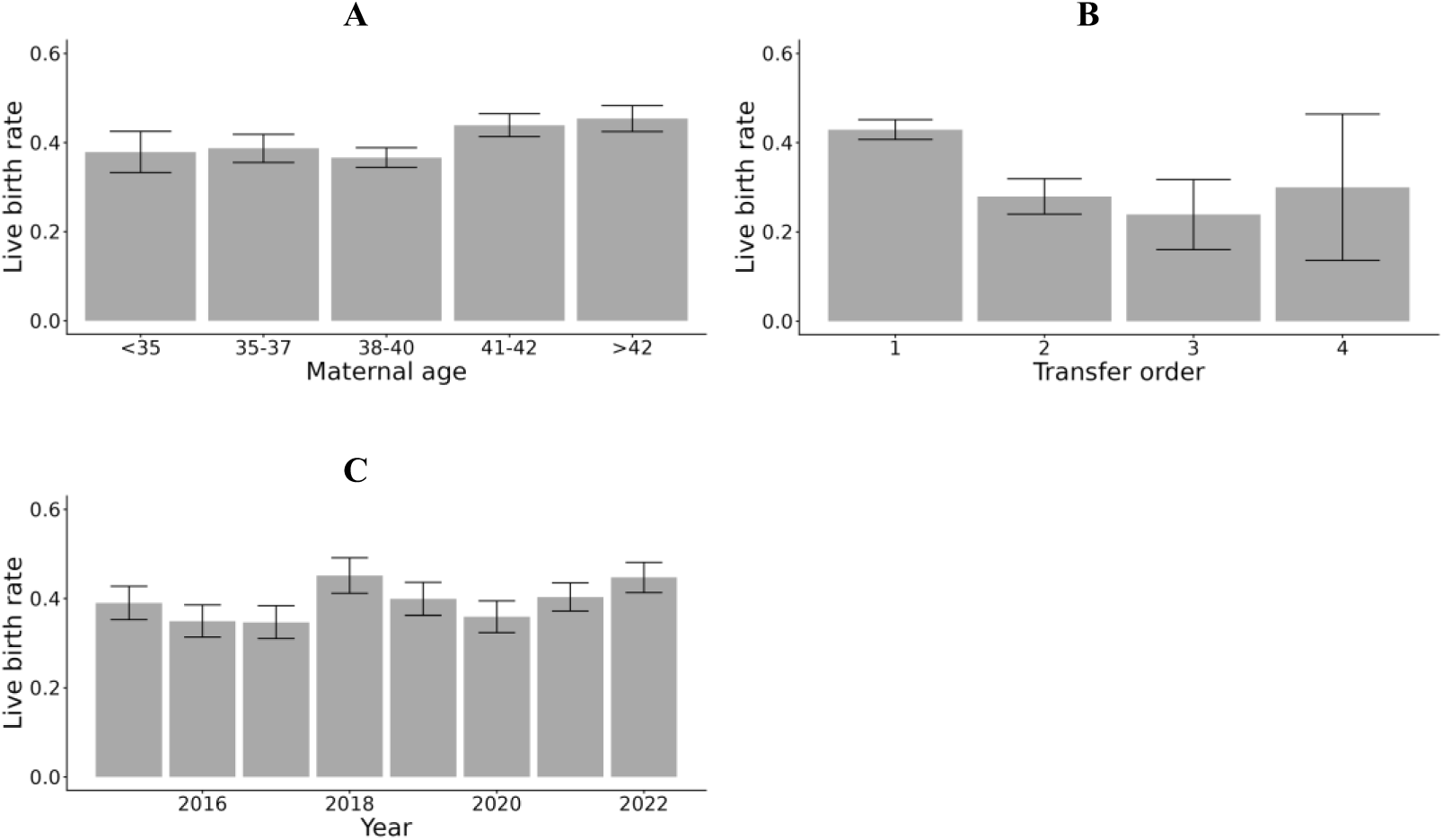
The live birth rate per euploid embryo transfer for cycles in which all embryos have been transferred. We show the average live birth rate per transfer vs the maternal age group (A), transfer number within the cycle (B), and year of treatment (C). The averages are over 5715 cycles. Bars denote 95% confidence intervals.

**Figure S4.**
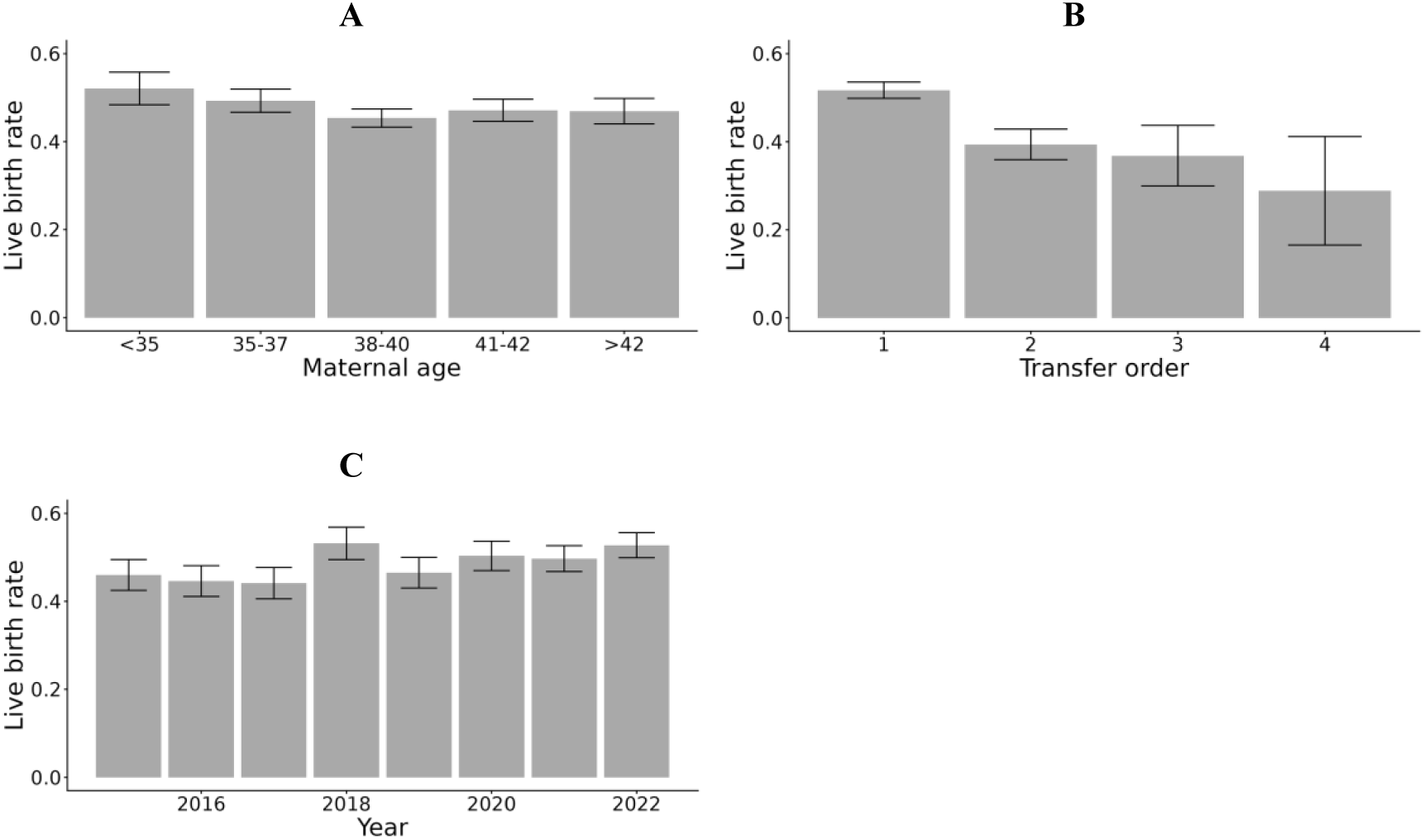
The live birth rate per euploid embryo transfer across all cycles. We show the average live birth rate per transfer vs the maternal age group (A), transfer number within the cycle (B), and year of treatment (C). The averages are over 6944 cycles. Bars denote 95% confidence intervals.

**Figure S5.**
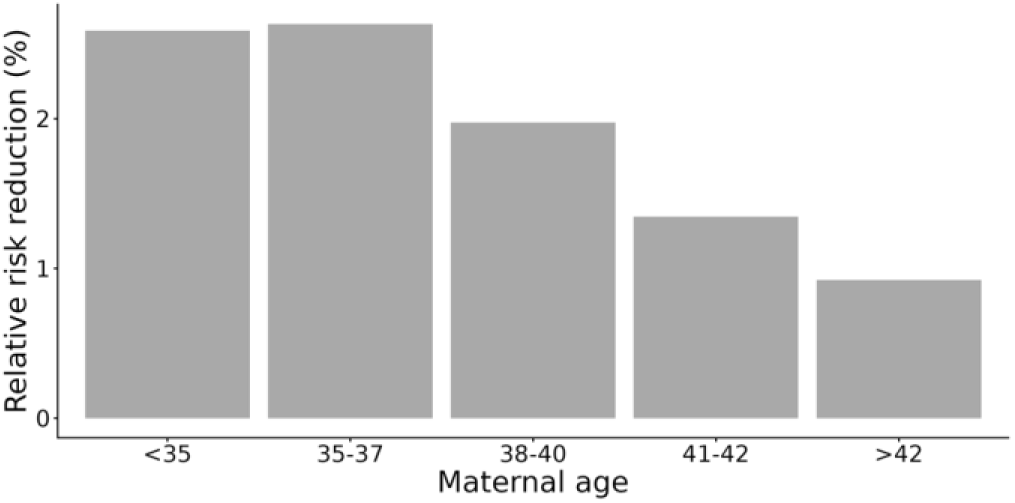
Risk reductions when transfer order is correlated with embryo PRS. The PRS accuracy was set to *r*^2^ = 0.1 and the disease prevalence to *K* = 0.01. We only used the 930 cycles in which all embryos have been transferred and that resulted in at least one live birth. The plot shows the RRR vs the maternal age group.

**Figure S6.**
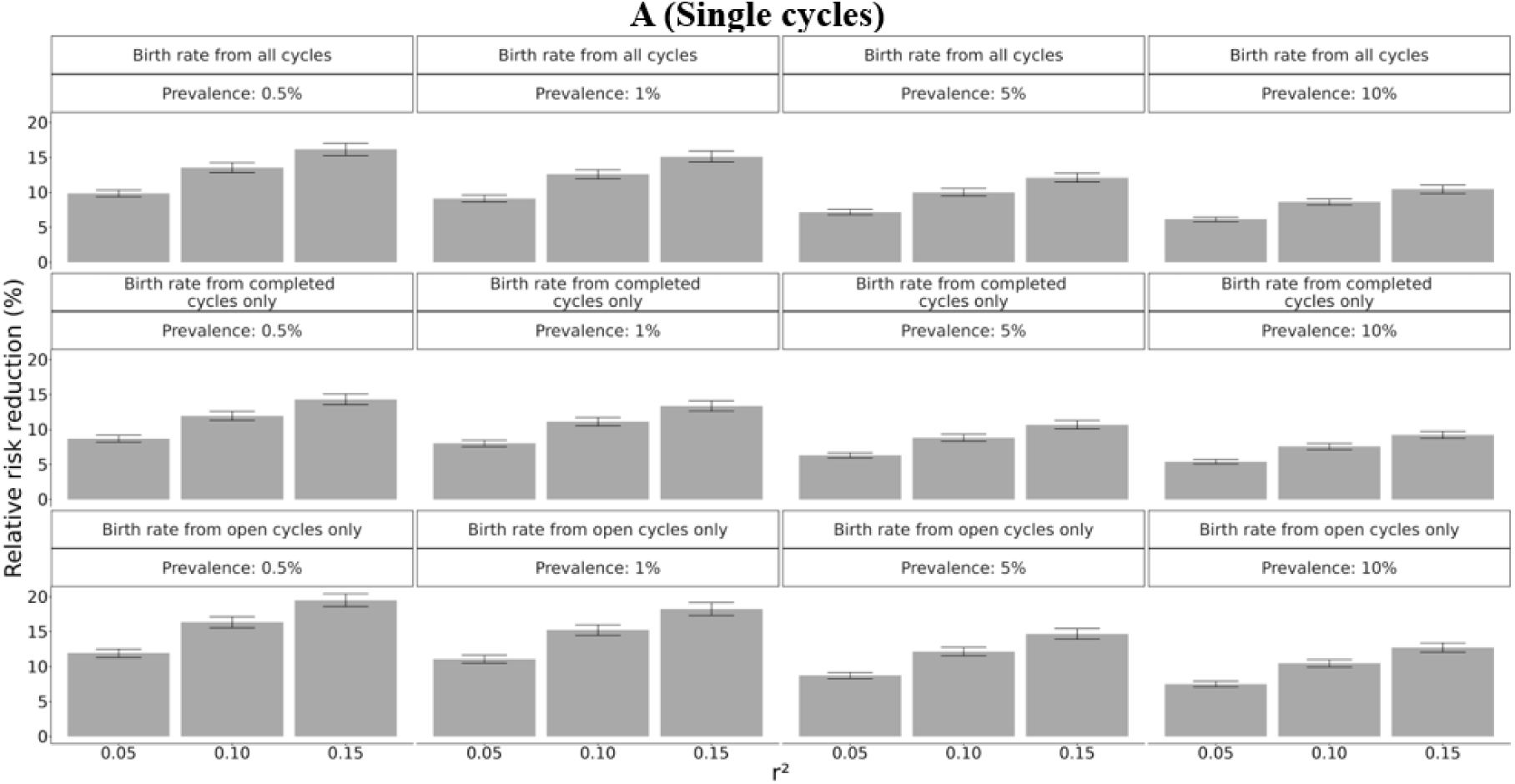
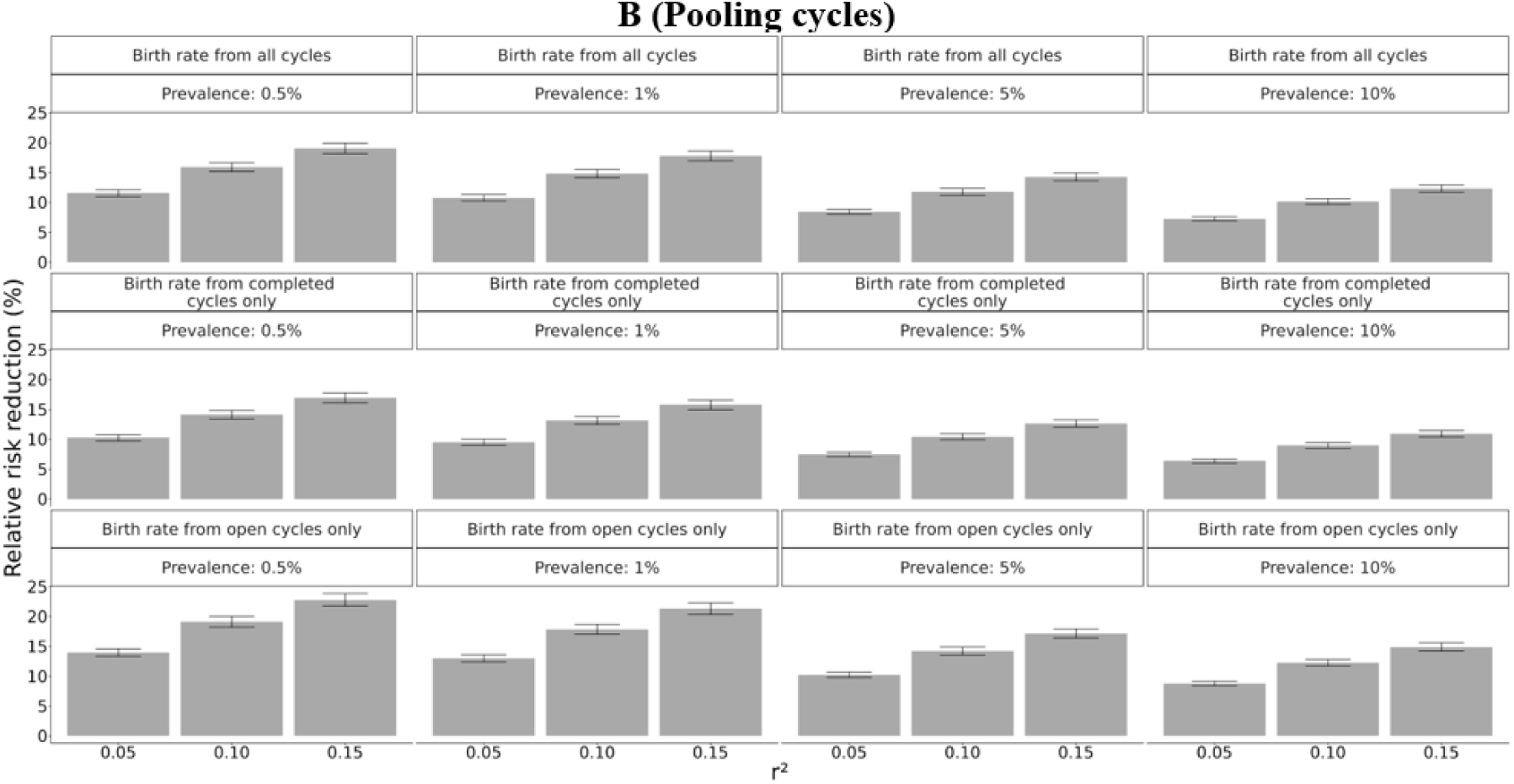
Sensitivity analyses in cycles with at least one birth. The figure is identical to Figure 5, except that we computed the risk reductions only in cycles with at least one birth. The analysis is based on the 930 cycles with at least one birth, as well as the 1229 open cycles. In the open cycles, we integrated over the outcomes of the non-transferred embryos, conditional on having at least one birth (Eq. (5)). Panel (A) corresponds to the outcomes of single cycle, while panel (B) uses pooled data from all cycles of the same couple.

**Figure S7.**
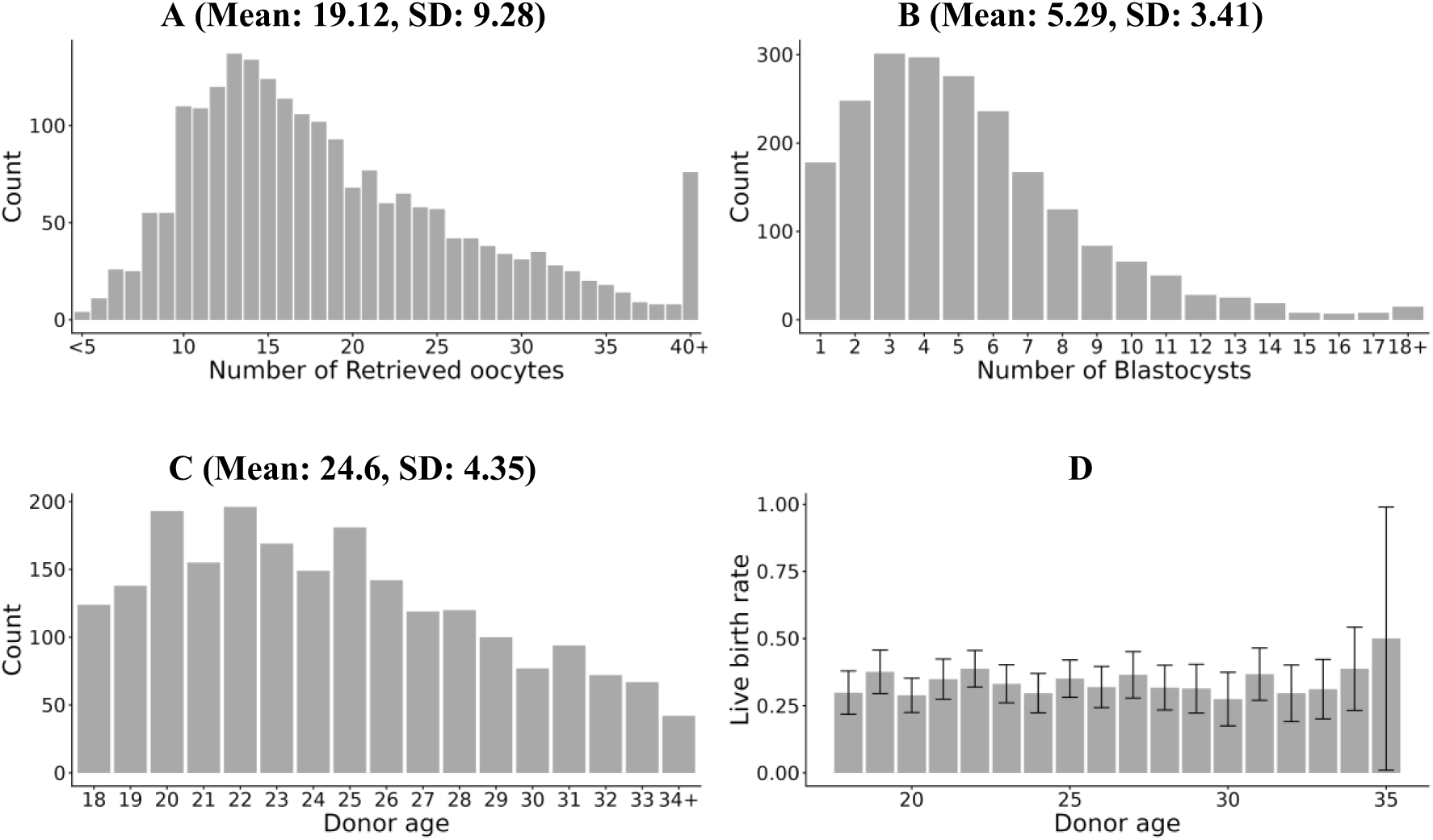
Properties of the egg donor cycles. (A) The distribution of the number of oocytes per cycle. Each donor appears in only one cycle. (B) The distribution of the number of blastocysts per cycle. (C) The distribution of the donor ages. (D) The live birth rate per blastocyst transfer vs the donor age (with 95% confidence intervals). We only considered cycles in which all oocytes were donated and all transfers were of single embryos.

**Figure S8.**
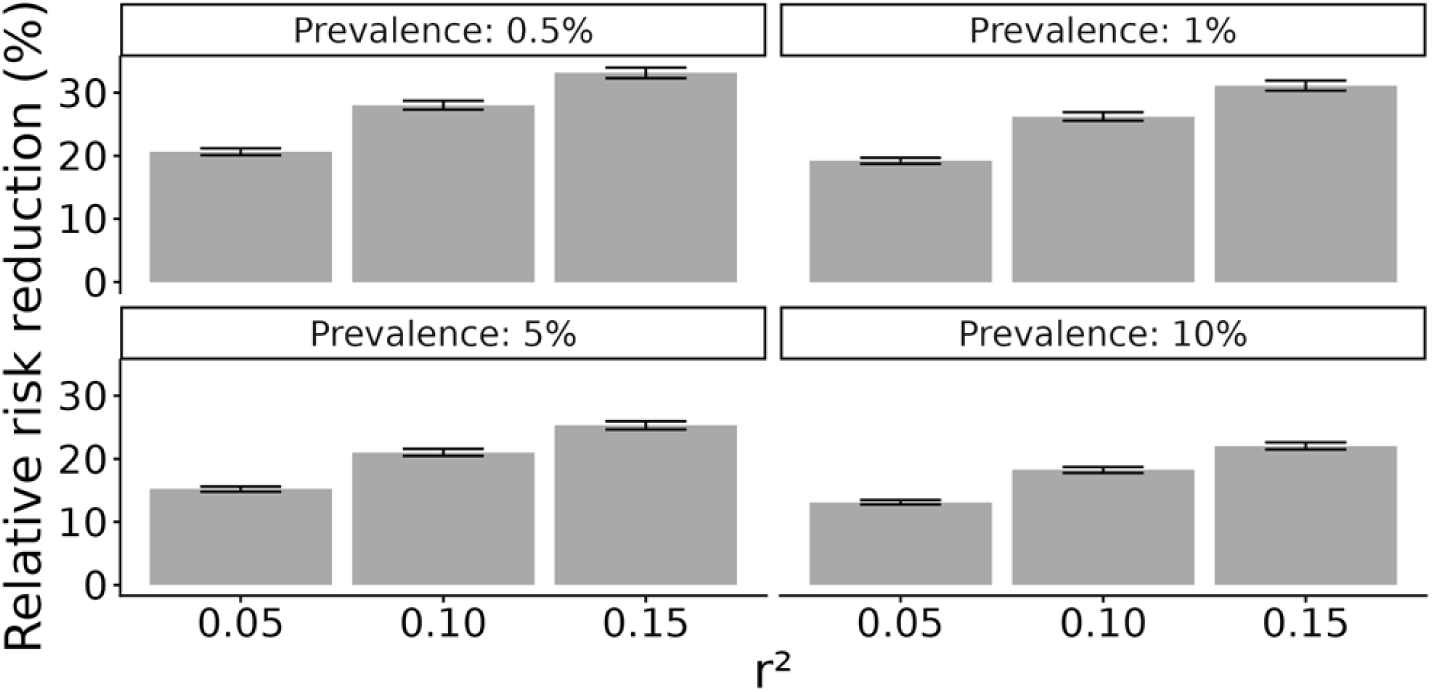
Risk reduction for egg donor cycles when conditioning on at least one live birth. The figure is exactly as in Figure 6, except that we condition on having at least one live birth per oocyte retrieval cycle. The risk reduction was computed using Eq. (5).

**Figure S9.**
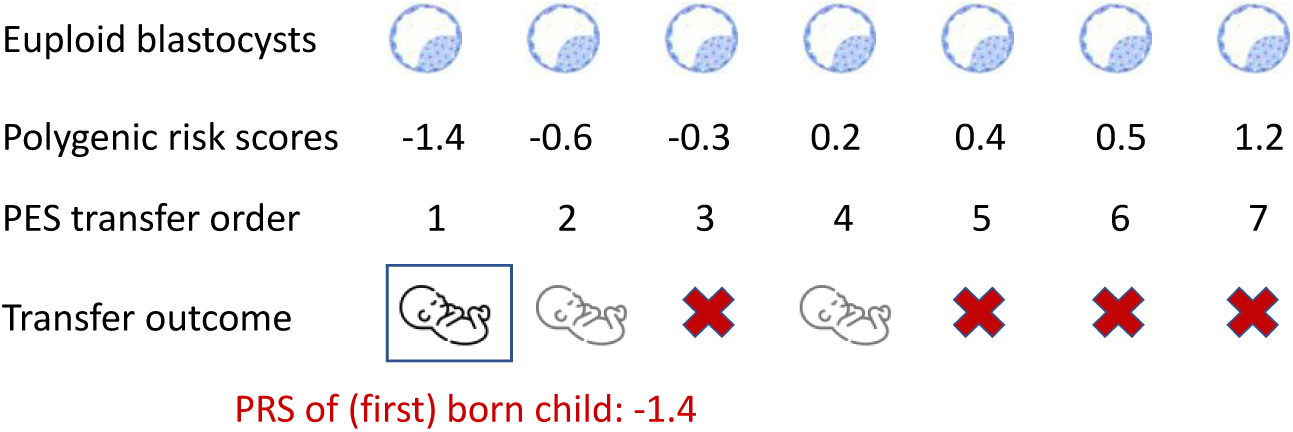
An illustration of embryo prioritization and the risk of the first-born child when transfer order is the same as ordering by PRS. As in Figure 1, we show the PRS for seven hypothetical embryos from the same cycle, all assumed to be euploid. Here, we assume that the real transfer order is identical to the ranking by PRS. In the example, because the first transfer was successful, the transfer of the lowest-PRS embryo results in a live birth, and the PRS of the first-born child is -1.4. This is lower than in Figure 1, where the first-born child had PRS -0.3. Generally, if the PRS order is correlated with the transfer order, we expect the first-born child to have lower risk compared to the case of no correlation. Such a correlation would also imply lower risk for the first-born child even without PES. However, to obtain an upper bound for the risk reduction due to PES, we compare the PES-based risk to that of the case of no PES and no correlation with transfer order.

## Notes

### Competing Interest Statement

EMP, FM, CSO, and AC are employees of Genethyx. Some of the work on this paper was performed when AC, FM, EMP, LP, and CSO were employees of Juno Genetics. SC is a paid consultant at MyHeritage.

### Funding Statement

The study was funded in part by the National Human Genome Research Institute of the National Institutes of Health under award number R01HG011711. The content is solely the responsibility of the authors and does not necessarily represent the official views of the National Institutes of Health.

### Author Declarations

For both datasets, approval was provided for the pseudonymous retrospective analysis of electronic medical records to test for the effect of IVF procedures, including embryo selection approaches, on clinical efficacy, efficiency and safety. The study of the Italian data (own eggs cycles) was approved by the Comitato Etico territoriale Lazio Area 1, protocol number 0747/2023. The study of the Spanish data (egg donor cycles) was approved by the CEIm-Hospital universitario y politecnico de La Fe, protocol number 2024-0405-1.

## References

Awadalla, M. S., Agarwal, R., Ho, J. R., McGinnis, L. K., & Ahmady, A. (2022). Effect of trophectoderm biopsy for PGT-A on live birth rate per embryo in good prognosis patients. Archives of Gynecology and Obstetrics, 306(4), 1321–1327. 10.1007/s00404-022-06679-x

Barlevy, D., Furrer, R. A., Kalapatapu, A., Martinez, A., Lencz, T., Carmi, S., Lázaro-Muñoz, G., & Pereira, S. (2025). Eugenics and polygenic embryo screening: Public, clinician, and patient perceptions of conditions versus traits. Genetics in Medicine, 27(9). 10.1016/j.gim.2025.101507

Capalbo, A., de Wert, G., Mertes, H., Klausner, L., Coonen, E., Spinella, F., Van de Velde, H., Viville, S., Sermon, K., Vermeulen, N., Lencz, T., & Carmi, S. (2024). Screening embryos for polygenic disease risk: A review of epidemiological, clinical, and ethical considerations. Human Reproduction Update, 30(5), 529–557. 10.1093/humupd/dmae012

Cimadomo, D., Rienzi, L., Conforti, A., Forman, E., Canosa, S., Innocenti, F., Poli, M., Hynes, J., Gemmell, L., Vaiarelli, A., Alviggi, C., Ubaldi, F. M., & Capalbo, A. (2023). Opening the black box: Why do euploid blastocysts fail to implant? A systematic review and meta-analysis. Human Reproduction Update, 29(5), 570–633. 10.1093/humupd/dmad010

Clarke, E. A., Thaler, J., Lazarov, V., Zerbib, A., Bayefsky, M., Gounko, D., Baird, M., Lee, J. A., & Copperman, A. B. (2025). Live birth outcomes after first donor oocyte embryo transfer: A comparison by treatment pathways in poor prognosis patients. Journal of Assisted Reproduction and Genetics, 42(12), 4339–4345. 10.1007/s10815-025-03708-x

Cordogan, S., Starr, D. B., Treff, N. R., Lanchbury, J., Goldstein, E., Sadeghi, K., Burmeister, M., Dayani, L., Macdonald, P., Keen-Kim, J. D., Fishel, S., Cervantes, E., & Folkersen, L. (2025). Within- and Between-Family Validation of Nine Polygenic Risk Scores Developed in 1.5 Million Individuals: Implications for IVF, Embryo Selection, and Reduction in Lifetime Disease Risk (p. 2025.10.24.25338613). medRxiv. 10.1101/2025.10.24.25338613

Demko, Z. P., Simon, A. L., McCoy, R. C., Petrov, D. A., & Rabinowitz, M. (2016). Effects of maternal age on euploidy rates in a large cohort of embryos analyzed with 24-chromosome single-nucleotide polymorphism–based preimplantation genetic screening. Fertility and Sterility, 105(5), 1307–1313. 10.1016/j.fertnstert.2016.01.025

Deng, Z., Tang, Z., Tan, Y., & Yang, Y. (2025). Exploring effective human egg donation policies: Global laws and experiences. Humanities and Social Sciences Communications, 12(1), 1404. 10.1057/s41599-025-05755-2

Dwoskin, E., & Torbati, Y. (2025, July 16). Inside the Silicon Valley push to breed super-babies. The Washington Post. https://www.washingtonpost.com/technology/2025/07/16/orchid-polygenic-screening-embryos-fertility/

Forzano, F., Antonova, O., Clarke, A., de Wert, G., Hentze, S., Jamshidi, Y., Moreau, Y., Perola, M., Prokopenko, I., Read, A., Reymond, A., Stefansdottir, V., van El, C., & Genuardi, M. (2022). The use of polygenic risk scores in pre-implantation genetic testing: An unproven, unethical practice. European Journal of Human Genetics, 30(5), Article 5. 10.1038/s41431-021-01000-x

Furrer, R. A., Barlevy, D., Pereira, S., Carmi, S., Lencz, T., & Lázaro-Muñoz, G. (2024). Public Attitudes, Interests, and Concerns Regarding Polygenic Embryo Screening. JAMA Network Open, 7(5), e2410832. 10.1001/jamanetworkopen.2024.10832

Goldman, R. H., Racowsky, C., Farland, L. V., Munne, S., Ribustello, L., & Fox, J. H. (2017). Predicting the likelihood of live birth for elective oocyte cryopreservation: A counseling tool for physicians and patients. Hum Reprod, 32(4), 853–859.

Grebe, T. A., Khushf, G., Greally, J. M., Turley, P., Foyouzi, N., Rabin-Havt, S., Berkman, B. E., Pope, K., Vatta, M., Kaur, S., & ACMG Social, Ethical, and Legal Issues Committee. (2024). Clinical utility of polygenic risk scores for embryo selection: A points to consider statement of the American College of Medical Genetics and Genomics (ACMG). Genetics in Medicine: Official Journal of the American College of Medical Genetics, 26(4), 101052. 10.1016/j.gim.2023.101052

Griffin, D. K., & Gordon, A. T. (2023). Preimplantation Testing for Polygenic Disease (PGT-P): Brave New World or Mad Pursuit? DNA, 3(2), Article 2. 10.3390/dna3020008

Huttler, A., Duvall, D., Sakkas, D., Heyward, Q., Sabbagh, R., Alper, M., & Vaughan, D. (2025). Achieving Two Live Births from One Ovarian Stimulation Cycle: The One-and-Done Approach Revisited. Fertility and Sterility. 10.1016/j.fertnstert.2025.07.029

Insogna, I. G., Lanes, A., Lee, M. S., Ginsburg, E. S., & Fox, J. H. (2021). Association of Fresh Embryo Transfers Compared With Cryopreserved-Thawed Embryo Transfers With Live Birth Rate Among Women Undergoing Assisted Reproduction Using Freshly Retrieved Donor Oocytes. JAMA, 325(2), 156–163. 10.1001/jama.2020.23718

Karavani, E., Zuk, O., Zeevi, D., Barzilai, N., Stefanis, N. C., Hatzimanolis, A., Smyrnis, N., Avramopoulos, D., Kruglyak, L., Atzmon, G., Lam, M., Lencz, T., & Carmi, S. (2019). Screening Human Embryos for Polygenic Traits Has Limited Utility. Cell, 179(6), 1424–1435 e8.

Klausner, L., Revital, A., Lencz, T., & Carmi, S. (2025). PEStimate: Predicting offspring disease risk after Polygenic Embryo Screening (p. 2025.09.05.25335168). medRxiv. 10.1101/2025.09.05.25335168

Klipstein, S., Abhari, S., Amato, P., Arjunan, A., Bakare, T., Bergman, K., Bayefsky, M., Beyhan, Z., Cameron, K., Crockin, S., Goldstein, J., Hyun, I., Kawwass, J., O’Brien, J., Comeaux Plowden, T., Quinn, G., Rebar, R., Robins, J., Shannon, C. N., … Trolice, M. (2026). Use of preimplantation genetic testing for polygenic disorders (PGT-P): An Ethics Committee opinion. Fertility and Sterility, 125(1), 24–30. 10.1016/j.fertnstert.2025.10.023

Kumar, A., Im, K., Banjevic, M., Ng, P. C., Tunstall, T., Garcia, G., Galhardo, L., Sun, J., Schaedel, O. N., Levy, B., Hongo, D., Kijacic, D., Kiehl, M., Tran, N. D., Klatsky, P. C., & Rabinowitz, M. (2022). Whole-genome risk prediction of common diseases in human preimplantation embryos. Nature Medicine, 28(3), 513–516. 10.1038/s41591-022-01735-0

Kushnir, V. A., Khanna, P., Barad, D. H., & Gleicher, N. (2014). Establishment of comparative performance criteria for IVF centers: Correlation of live birth rates in autologous and donor oocyte IVF cycles. Reproductive Biology and Endocrinology, 12(1), 122. 10.1186/1477-7827-12-122

La Marca, A., Capuzzo, M., Longo, M., Imbrogno, M. G., Spedicato, G. A., Fiorentino, F., Spinella, F., Greco, P., Minasi, M. G., & Greco, E. (2022). The number and rate of euploid blastocysts in women undergoing IVF/ICSI cycles are strongly dependent on ovarian reserve and female age. Human Reproduction, 37(10), 2392–2401. 10.1093/humrep/deac191

Lazaro-Munoz, G., Pereira, S., Carmi, S., & Lencz, T. (2021). Screening embryos for polygenic conditions and traits: Ethical considerations for an emerging technology. Genet Med, 23(3), 432–434.

Lencz, T., Backenroth, D., Granot-Hershkovitz, E., Green, A., Gettler, K., Cho, J. H., Weissbrod, O., Zuk, O., & Carmi, S. (2021). Utility of polygenic embryo screening for disease depends on the selection strategy. eLife, 10, e64716.

Lencz, T., Bhattacharyya, U., Klausner, L., John, J., & Carmi, S. (2026). Clinical implications of rare and common variation in preimplantation genetic testing for breast cancer. Npj Genomic Medicine, 11(1), 4. 10.1038/s41525-025-00546-9

Lencz, T., Sabatello, M., Docherty, A., Peterson, R. E., Soda, T., Austin, J., Bierut, L., Crepaz-Keay, D., Curtis, D., Degenhardt, F., Huckins, L., Lazaro-Munoz, G., Mattheisen, M., Meiser, B., Peay, H., Rietschel, M., Walss-Bass, C., & Davis, L. K. (2022). Concerns about the use of polygenic embryo screening for psychiatric and cognitive traits. The Lancet Psychiatry, 9(10), 838–844. 10.1016/S2215-0366(22)00157-2

Li, S., Giardina, T., Katz, M., Chandramohan, D., Slotnick, N., Behr, B., Siddiqui, N., Xia, Y., & Podgursky, B. (2024). Concordance of whole-genome amplified embryonic DNA with the subsequently born child (p. 2024.01.12.24301086). medRxiv. 10.1101/2024.01.12.24301086

Lindner, P., Flannagan, K., Li, H. J., Wang, J., Hoyos, L. R., Uhler, M. L., Homer, M., Devine, K., Hill, M., & Romanski, P. (2025). Live birth outcomes after euploid transfer: Autologous vs. donor oocyte embryos in patients aged > 35 years. F&S Reports, 6(4), 462–469. 10.1016/j.xfre.2025.08.006

Marshall, L. (2017). Ethical Issues in the Evolving Realm of Egg Donation. In L. Francis (Ed.), The Oxford Handbook of Reproductive Ethics (p. 0). Oxford University Press. 10.1093/oxfordhb/9780199981878.013.21

Moore, S., Davidson, I., Anomaly, J., Li, J. H., Ahangari, M., Moissiy, L., Christensen, M., Young, A. S., Stern, D., & Wolfram, T. (2025). Development and validation of polygenic scores for within-family prediction of disease risks (p. 2025.08.06.25333145). medRxiv. 10.1101/2025.08.06.25333145

Nielsen, N. M., Westergaard, T., Rostgaard, K., Frisch, M., Hjalgrim, H., Wohlfahrt, J., Koch-Henriksen, N., & Melbye, M. (2005). Familial Risk of Multiple Sclerosis: A Nationwide Cohort Study. American Journal of Epidemiology, 162(8), 774–778. 10.1093/aje/kwi280

Pereira, S., Carmi, S., Altarescu, G., Austin, J., Barlevy, D., Hershlag, A., Juengst, E., Kostick-Quenet, K., Kovanci, E., Lathi, R. B., Mukherjee, M., Van den Veyver, I., Zuk, O., Lázaro-Muñoz, G., & Lencz, T. (2022). Polygenic embryo screening: Four clinical considerations warrant further attention. Human Reproduction, 37(7), 1375–1378. 10.1093/humrep/deac110

Polyakov, A., Amor, D. J., Savulescu, J., Gyngell, C., Georgiou, E. X., Ross, V., Mizrachi, Y., & Rozen, G. (2022). Polygenic risk score for embryo selection—Not ready for prime time. Human Reproduction, 37(10), 2229–2236. 10.1093/humrep/deac159

Rodríguez-Varela, C., Mascarós, J. M., Labarta, E., Silla, N., & Bosch, E. (2024). Minimum number of mature oocytes needed to obtain at least one euploid blastocyst according to female age in in vitro fertilization treatment cycles. Fertility and Sterility, 122(4), 658–666. 10.1016/j.fertnstert.2024.06.002

Shams, H., Shao, X., Santaniello, A., Kirkish, G., Harroud, A., Ma, Q., Isobe, N., University of California San Francisco MS-EPIC Team, Schaefer, C. A., McCauley, J. L., Cree, B. A. C., Didonna, A., Baranzini, S. E., Patsopoulos, N. A., Hauser, S. L., Barcellos, L. F., Henry, R. G., & Oksenberg, J. R. (2023). Polygenic risk score association with multiple sclerosis susceptibility and phenotype in Europeans. Brain, 146(2), 645–656. 10.1093/brain/awac092

Siermann, M., Valcke, O., Vermeesch, J. R., Raivio, T., Tšuiko, O., & Borry, P. (2024). “Are we not going too far?“: Socio-ethical considerations of preimplantation genetic testing using polygenic risk scores according to healthcare professionals. Social Science & Medicine, 343, 116599. 10.1016/j.socscimed.2024.116599

Siermann, M., Vermeesch, J. R., Raivio, T., Tšuiko, O., & Borry, P. (2024). Polygenic embryo screening: Quo vadis? Journal of Assisted Reproduction and Genetics, 41(7), 1719–1726. 10.1007/s10815-024-03169-8

Sud, A., Horton, R. H., Hingorani, A. D., Tzoulaki, I., Turnbull, C., Houlston, R. S., & Lucassen, A. (2023). Realistic expectations are key to realising the benefits of polygenic scores. BMJ, 380, e073149. 10.1136/bmj-2022-073149

Tiegs, A. W., Tao, X., Zhan, Y., Whitehead, C., Kim, J., Hanson, B., Osman, E., Kim, T. J., Patounakis, G., Gutmann, J., Castelbaum, A., Seli, E., Jalas, C., & Scott, R. T. (2021). A multicenter, prospective, blinded, nonselection study evaluating the predictive value of an aneuploid diagnosis using a targeted next-generation sequencing–based preimplantation genetic testing for aneuploidy assay and impact of biopsy. Fertility and Sterility, 115(3), 627–637. 10.1016/j.fertnstert.2020.07.052

Treff, N. R., Eccles, J., Lello, L., Bechor, E., Hsu, J., Plunkett, K., Zimmerman, R., Rana, B., Samoilenko, A., Hsu, S., & Tellier, L. (2019). Utility and First Clinical Application of Screening Embryos for Polygenic Disease Risk Reduction. Front Endocrinol (Lausanne), 10, 845.

Treff, N. R., Eccles, J., Marin, D., Messick, E., Lello, L., Gerber, J., Xu, J., & Tellier, L. (2020). Preimplantation Genetic Testing for Polygenic Disease Relative Risk Reduction: Evaluation of Genomic Index Performance in 11,883 Adult Sibling Pairs. Genes (Basel), 11(6). https://www.ncbi.nlm.nih.gov/pubmed/32545548

Treff, N. R., Savulescu, J., de Melo-Martín, I., Shulman, L. P., & Feinberg, E. C. (2022). Should preimplantation genetic testing for polygenic disease be offered to all—Or none? Fertility and Sterility, 117(6), 1162–1167. 10.1016/j.fertnstert.2022.03.017

Treff, N. R., Zimmerman, R., Bechor, E., Hsu, J., Rana, B., Jensen, J., Li, J., Samoilenko, A., Mowrey, W., Van Alstine, J., Leondires, M., Miller, K., Paganetti, E., Lello, L., Avery, S., Hsu, S., & Melchior Tellier, L. C. A. (2019). Validation of concurrent preimplantation genetic testing for polygenic and monogenic disorders, structural rearrangements, and whole and segmental chromosome aneuploidy with a single universal platform. Eur J Med Genet.

Turley, P., Meyer, M. N., Wang, N., Cesarini, D., Hammonds, E., Martin, A. R., Neale, B. M., Rehm, H. L., Wilkins-Haug, L., Benjamin, D. J., Hyman, S., Laibson, D., & Visscher, P. M. (2021). Problems with Using Polygenic Scores to Select Embryos. N Engl J Med, 385(1), 78–86.

Vergallo, G. M., Marinelli, S., Napoletano, G., Paola, L. D., Treglia, M., Zaami, S., & Frati, P. (2025). 20 Years Since the Enactment of Italian Law No. 40/2004 on Medically Assisted Procreation: How It Has Changed and How It Could Change. International Journal of Environmental Research and Public Health, 22(2). 10.3390/ijerph22020296

Wald, N. J., & Old, R. (2019). The illusion of polygenic disease risk prediction. Genet Med, 21(8), 1705–1707.

Widen, E., Lello, L., Raben, T. G., Tellier, L. C. A. M., & Hsu, S. D. H. (2022). Polygenic Health Index, General Health, and Pleiotropy: Sibling Analysis and Disease Risk Reduction. Scientific Reports, 12(1), 18173. 10.1038/s41598-022-22637-8

